# Safety, harm, and efficacy of IRRAflow^®^ versus external ventricular drainage for intraventricular hemorrhage: A randomized clinical trial

**DOI:** 10.1101/2023.07.08.23292286

**Authors:** Mette Haldrup, Mads Rasmussen, Niwar Mohamad, Stig Dyrskog, Line Thorup, Nikola Mikic, Joakim Wismann, Mads Grønhøj, Frantz Rom Poulsen, Mojtaba Nazari, Naveed Ur Rehman, Claus Ziegler Simonsen, Anders Rosendal Korshøj

## Abstract

**Importance:** Intraventricular hemorrhage (IVH) is associated with high morbidity and mortality. A strong need exists for treatment advances. IRRAflow^®^ was recently introduced as a method for minimally invasive, controlled, and accelerated IVH wash-out. However, no current evidence supports this technology. Here, we present the first pivotal safety/efficacy evaluation in a randomized controlled setting.

**Objective:** To evaluate the safety and efficacy of active hematoma irrigation using the IRRAflow^®^ device compared with standard external ventricular drainage (EVD) for the IVH treatment.

**Design, setting, and participants:** This investigator-initiated, prospective multi-center, 1:1 randomized, single-blinded, clinical trial was conducted at the Aarhus University Hospital and Odense University Hospital in Denmark from January 13, 2022 to November 24, 2022. The trial was set to include 58 IVH patients with prespecified interim analysis at final endpoint collection for the first 20 patients.

**Interventions:** Patients were randomized to receive either IRRAflow^®^ or standard EVD treatment. The IRRAflow^®^ performs periodic active irrigation and aspiration contrary to standard passive gravity-driven EVD.

**Main outcomes and measures:** Outcomes were chosen to reflect key IRRAflow^®^ value propositions. The primary outcome was rate of catheter occlusions. The main outcome of the prespecified interim analysis was risk of severe adverse events (SAEs). Secondary outcomes were hematoma clearance rate, shunt dependency rate, procedure time for primary catheter placement, length of intensive care unit (ICU) stay, treatment duration, functional outcome (modified Rankin Scale (mRS) and extended Glasgow Outcome Scale (eGOS)) 90 days after inclusion, adverse events (AEs) (including catheter-related infections and procedure-related complications), and overall survival.

**Results:** The study was terminated early due to a significantly increased risk of SAEs in the IRRAflow^®^ group compared with EVD (risk difference = 0.43, 95% confidence interval (CI) (0.059;0.813), p=0.044), incidence rate ratio divided by person time = 6.0, 95%CI 1.38-26.11) p=0.012).

The catheter occlusion rate was 37.5% in the IRRAflow^®^ group versus 6.6% in the EVD group (p=0.141), which met the prespecified 0.2 alpha level. The median procedure time for primary catheter placement was 53.5 min compared with 12 min in the control group (p=0.0001). No significant difference was observed for other secondary outcomes. The majority of SAEs had a causal relationship with the intervention.

**Conclusion and relevance:** We found that the current IRRAflow^®^ treatment is encumbered by a significantly increased number of SAEs. We recommend caution when using the device. Based on root-cause analysis, our recommendation is that a number of changes be implemented to improve the safety of the device in IVH treatment. We believe that our results are pivotal in ensuring the future safety of patients with IVH.

**Trial registration:** ClicalTrials.gov identifier: NCT05204849, registered 15 December 2021, updated 24 December 2022.

## Introduction

Intraventricular hemorrhage (IVH) is a serious hemorrhagic stroke subtype characterized by bleeding within the cerebral ventricles. IVH is associated with high mortality rates of up to 50% and causes severe disability in survivors^1^. Current treatment relies on supportive care and cerebrospinal fluid (CSF) drainage to decompress the intracranial space and facilitate passive hematoma evacuation using an external ventricular drain (EVD). Recent studies show that intraventricular fibrinolysis (IVF) may accelerate clot removal, increasing overall survival and improving functional outcomes. It was suggested that further acceleration of clot removal is required to significantly improve patient outcome^2,3^. However, recent decades have seen no serious treatment advances and minimally invasive techniques aimed at this goal have been limited.

Recently, a new technology coined IRRAflow^®^ (IRRAS AB) was introduced to address unmet clinical needs in IVH treatment. IRRAflow^®^ uses a dual-lumen catheter to perfuse the ventricular system with physiological saline while monitoring intracranial pressure (ICP). This continuous irrigation and aspiration is hypothesized to accelerate IVH wash-out and to prevent complications like catheter occlusion and bacterial infection commonly associated with passive drainage. IRRAflow^®^ received CE marketing and FDA approval in 2019 and 2018, respectively, and has been used to treat more than 600 patients with IVH, brain abscess, and chronic subdural hematoma worldwide (post-market surveillance data, courtesy of IRRAS AB). Currently, however, no evidence supports the safety or efficacy of such treatment. Only one published case report (DRIFT) has evaluated IRRAflow^®^ for IVH treatment^4^, finding indications of accelerated IVH clearance without adverse events (AEs). In addition, one registered randomized controlled trial (ARCH (NCT05118997) is currently evaluating IRRAflow^®^ in combination with IVF for patients with IVH. Furthermore, three unregistered trials (VASH, AFFECT, and DIVE) are presently underway testing IRRAflow^®^ in various settings.

In this study (ACTIVE - Active fluid exchange in the treatment of IVH, NCT05204849), we present the safety and preliminary efficacy of IRRAflow^®^ against standard EVD for IVH treatment. The study aimed to investigate the major value propositions of the technology, particularly reduced rates of drain occlusion and central nervous system (CNS) infection, and accelerated hematoma clearance and. Presenting first class-1 evidence on IRRAflow^®^ performance and operation under basic recommended treatment conditions, this study provides an important basis for future studies and developments in this area.

## Methods

### Trial design and oversight

ACTIVE is a single-blinded, phase-two trial evaluating the safety and efficacy of the IRRAflow^®^ system versus passive EVDs for treating IVH. It is a prospective, investigator-initiated, multi-center, randomized, comparative trial conducted at two Danish university hospitals (Aarhus (Sponsor) and Odense). The study abides by the ethical principles of the Declaration of Helsinki, Good Clinical Practice Guidelines, and ISO-14155 standards, with approval from the Danish Central Region Committee on Health Research Ethics. Informed consent was obtained and the trial was monitored by an independent data safety monitoring committee (DMC) and the contract research organization. The study followed the Consolidated Standards of Reporting Trials (CONSORT) reporting guideline, with harms reported following the CONSORT extension for harm reporting^5^. Details of trial design, conduct, oversight, and analysis are provided in the study protocol (Supplementary Material 1 and Haldrup et al. 2022^6^). The trial was registered with ClinicalTrials.gov (NCT05204849) on December 15, 2021 and updated on January 1, 2022. A prespecified interim analysis was planned after three months of follow-up for the first 20 participants.

### Participants

Patients aged ≥ 18 years with primary or secondary IVH and indication for CSF drainage were assessed for eligibility. An IVH Graeb score > 3 was required for inclusion (computed tomography (CT) required < 24 hours before inclusion). Patients with fixed and dilated pupils were excluded as were pregnant and nursing women. A full list of inclusion and exclusion criteria is provided in the study protocol.

### Randomization and blinding

Participants were randomly assigned 1:1 for treatment with IRRAflow (intervention) or standard EVD (control), with randomization performed by the coordinating investigator upon patient enrollment using the REDCap randomization module. The allocation sequence was provided by the REDCap administrator at Aarhus University. Patients received a patient ID upon enrollment. The patient ID was not known to anyone outside of the research group.

To guarantee study integrity, participants were blinded to randomization and treatment, whereas the independent study statistician was blinded to endpoint assessment. The independent DMC had full source data access upon request and unblinding privileges for internal safety assessments.

### Interventions

Intervention group participants were treated with IRRAflow^®^ (dual-lumen catheter, version 2.0 ICGS020, using control unit, version 3.0). Control group participants were treated with a standard EVD (Medtronic Inc., Silverline^®^, size f10) that was bolted and silver impregnated. All surgeries were conducted or supervised by trained neurosurgeons, and appropriate surgical guidelines and technical product recommendations were followed. Catheters were placed using neuronavigation guidance (Medtronic, Axiem) and positioned in the lateral ventricle with the least amount of blood needed to increase the likelihood of maintaining perioperative catheter drainage integrity. The catheter’s tip and draining part were positioned in the CSF and outside the hematoma to avoid obstruction.

In case of obstruction of the foramen Monro, a standard EVD (Medtronic Inc., Silverline^®^) was placed contralaterally to ensure bilateral drainage. The IRRAflow^®^ device was set to one of three basic irrigation modes (low - 20 ml/hr, medium - 90 ml/hr, and high 180 ml/hr); modes were continuously adjusted by the treating physician to maximize hematoma irrigation while maintaining patient safety (see Supplementary 2 and 3). All participants treated with IRRAflow^®^ had an ipsilateral Raumedic Neurovent-P parenchymal ICP sensor to validate their IRRAflow^®^ pressure measurements.

Study personnel were thoroughly trained in all study procedures, including use and operation of IRRAflow^®^, which was provided by a qualified IRRAS associate or designee.

### Primary outcome

The primary outcome was rate of catheter occlusion defined as complete drainage stop. This outcome was chosen to test the performance of IRRAflow^®^ on one of its key hypothesized value propositions.

### Safety outcomes

During the prespecified interim analysis, the safety outcome was the risk of severe adverse events (SAEs). If the risk of SAEs associated with IRRAflow^®^ was increased compared with EVD at the 0.20 significance level (Poisson regression), early discontinuation would be considered.

Adverse events (AEs) were defined as any unfavorable and unintended sign, symptom, or disease associated with use of a medical treatment or procedure. AE severity was graded according to the Common Terminology Criteria of Adverse Events (CTCAE) version 5.0 with AEs grade 3 to 5 categorized as SAEs. All AEs and SAEs were recorded and their causality evaluated using the WHO-UMC system for standardized case causality assessment^7^. AEs certainly, probably, or possibly related to the intervention or catheter treatment were analyzed. In case of recurrent AEs, the risk of experiencing > 1 AE was analyzed along with the incidence of all AEs in the two groups (Supplementary Material 4).

All SAEs were evaluated thoroughly in a root cause analysis including individual review and validation by the independent DMC. The independent neurosurgical DMC member determined the causal relationship of all SAEs with EVD treatment in general and the specific feature differences between IRRAflow^®^ and EVD (Figure 1, Supplementary Material 2). All endpoints, including AEs, were prospectively registered during the 90-day follow-up period.

**Figure 1.**
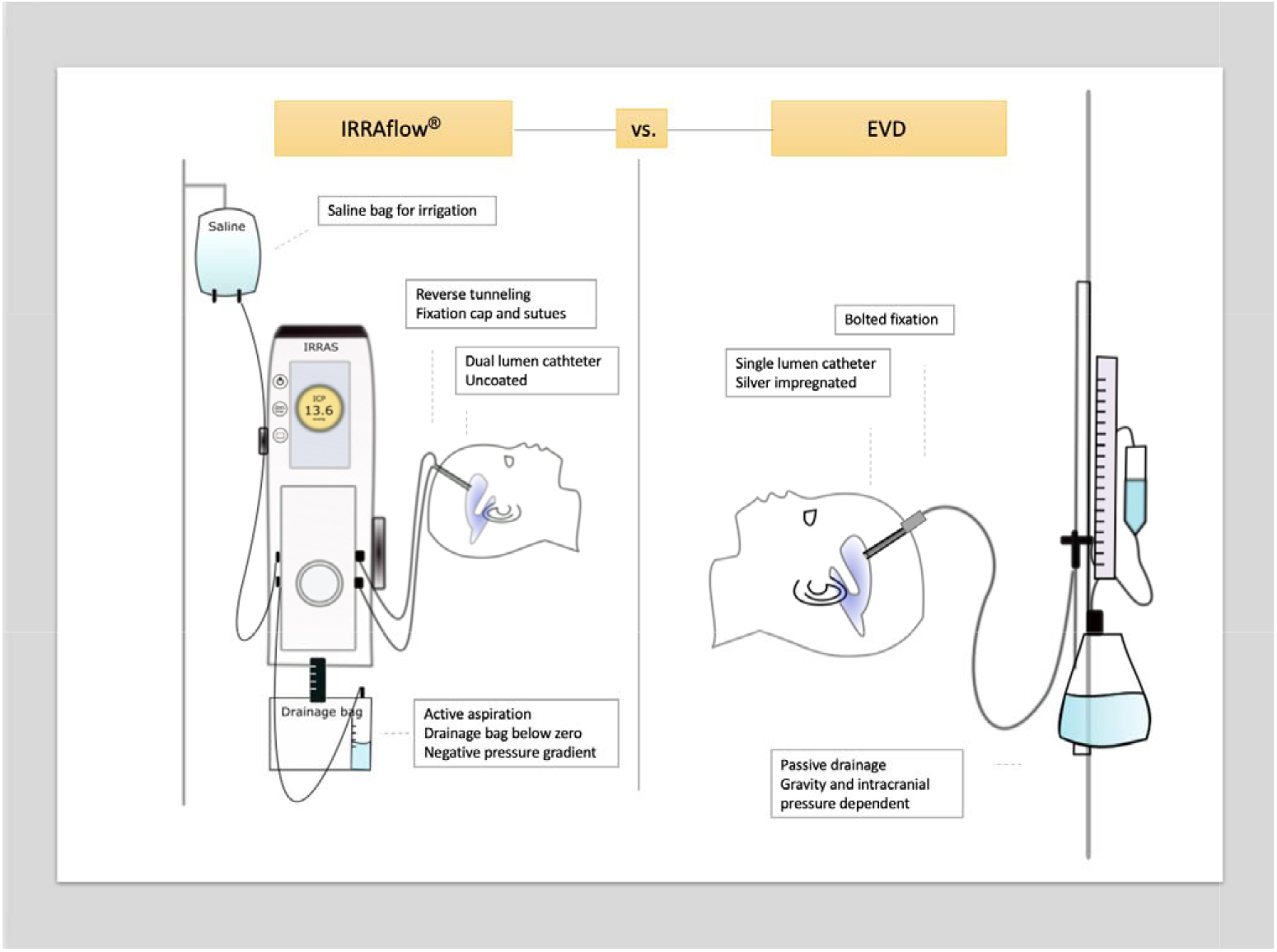
The IRRAflow^®^ device (intervention) and standard passive EVD (control). Differences between the IRRAflow^®^ and the EVD are outlined in the boxes.

### Secondary outcomes

The final analysis assessed the following secondary outcomes: (1) proportion of participants with an implanted CSF shunting device at 90 days, (2) IVH clearance rate based on CT volumetrics at days 0, 2, 4, 6, and 8 after inclusion, (3) procedure time for primary catheter placement, (4) length of ICU stay, (5) catheter treatment duration, (6) functional outcome using the modified Rankin Scale (mRS) score at inclusion, discharge, and 90 days after inclusion, and extended Glasgow Outcome Scale (eGOS) at discharge and 90 days, (7) AEs including catheter-related infections and procedure-related complications, and (8) overall survival. These outcomes were chosen to reflect additional IRRAflow^®^ value propositions, clinical feasibility and importance.

### Sample size calculation

The study was designed to enroll 58 participants based on an assumption of a superiority effect in the primary outcome of catheter occlusions. We used a two-sided log-rank test to compare the time-to-catheter occlusion between the two treatment groups, assuming an equal length of follow-up and an occlusion risk of 10% and 35% in the intervention and control groups, respectively. We set a more conservative occlusion rate estimate than reported in the literature for the control group and a higher occlusion risk than the 0% reported in post-market surveillance by IRRAS AB1,8,9. Patient mortality was assumed to be independent of the time to catheter occlusion and to correspond to a 30% dropout rate. To achieve a power of 80% using Schoenfeld’s approach, we required a sample size of 58 (i.e. 29 subjects with a drain per treatment group). The study was exploratory in nature, and we used a 20% significance level to take this into account.

### Statistical analysis

The null-hypothesis was that active irrigation has no beneficial effect on the occlusion rate compared with standard passive EVD.

Final intention-to-treat analyses were conducted based on the full study population for all endpoints, including safety. Per-protocol analyses were conducted as relevant. We performed statistical tests at a two-sided significance level of 0.05, except for primary outcomes of interim and final analyses, which were set to 0.2.

Time-to-catheter occlusion was analyzed for the primary catheter alone. As some participants were treated with bilateral catheters, analysis was also conducted for all collective primary and secondary catheters using plots of cumulative incidence ratios and Cox regression. Other time-to-event endpoints were analyzed with Kaplan-Meier estimates and Cox regression. Catheter-related infections and shunt dependency were reported as proportions using a generalized linear model with log-link function, risk ratios, 95% confidence intervals (CI) intervals, and p-values. Length of ICU stay was evaluated using Student’s t-test and reported as means (+/-SE). Ventricular blood clearance was analyzed using a linear mixed effect regression model. Functional outcome measures (mRS and eGOS) were dichotomized and analyzed using baseline-adjusted logistic regression. All estimates were reported with corresponding CIs. Procedure time and catheter treatment duration were reported using median estimates and Bonferroni-adjusted CIs. Participants were censored upon death. SAEs were analyzed as absolute risk differences and incidence rates divided by person-time to account for each participant’s variable time contribution. Incidence rate ratios were estimated using the Poisson regression model.

## Results

Twenty-four participants were assessed for eligibility between January 13, 2022, and November 24, 2022. Among these, 21 participants (11 intervention and 10 control) met all inclusion criteria and were included. Their baseline characteristics are shown in Table 1. No statistical difference was observed between the two groups’ baseline characteristics. The participant flow chart is displayed in Figure 2. The trial’s final endpoint was collected on February 14, 2023.

**Table 1.** Baseline characteristics of the 21 participants included in the ACTIVE Study, 11 participants in the intervention group and 10 participants in the control group.

|  | Intervention | Control | p-value |
| --- | --- | --- | --- |
| Age, median (range) | 66 (41-86) | 68.5 (42-81) | 0.899 |
| Male (n) | 8 | 6 | 0.659 |
| Female (n) | 3 | 4 |  |
| Antihypertensive medicine compliant (%) | 27.3% | 60% | 0.131 |
| Anticoagulated at enrollment (%) | 9.1% | 20% | 0.476 |
| Antiplatelet treatment at enrollment (%) | 27.3% | 10% | 0.313 |
| IVH type |  |  |  |
| SAH with intraventricular extension (n) | 6 | 4 | 0.520 |
| ICH with intraventricular extension (n) | 4 | 6 |  |
| Primary IVH (n) | 1 | 0 |  |
| GCS at inclusion, median (range) | 6 (3-12) | 7 (3-13) | 0.812 |
| Graeb score, median (range) | 8 (3-11) | 9.5 (6-12) | 0.07 |

IVH = Intraventricular hemorrhage
SAH = Subarachnoid hemorrhage
ICH = Intracerebral hemorrhage
GCS = Glasgow coma scale

**Figure 2.**
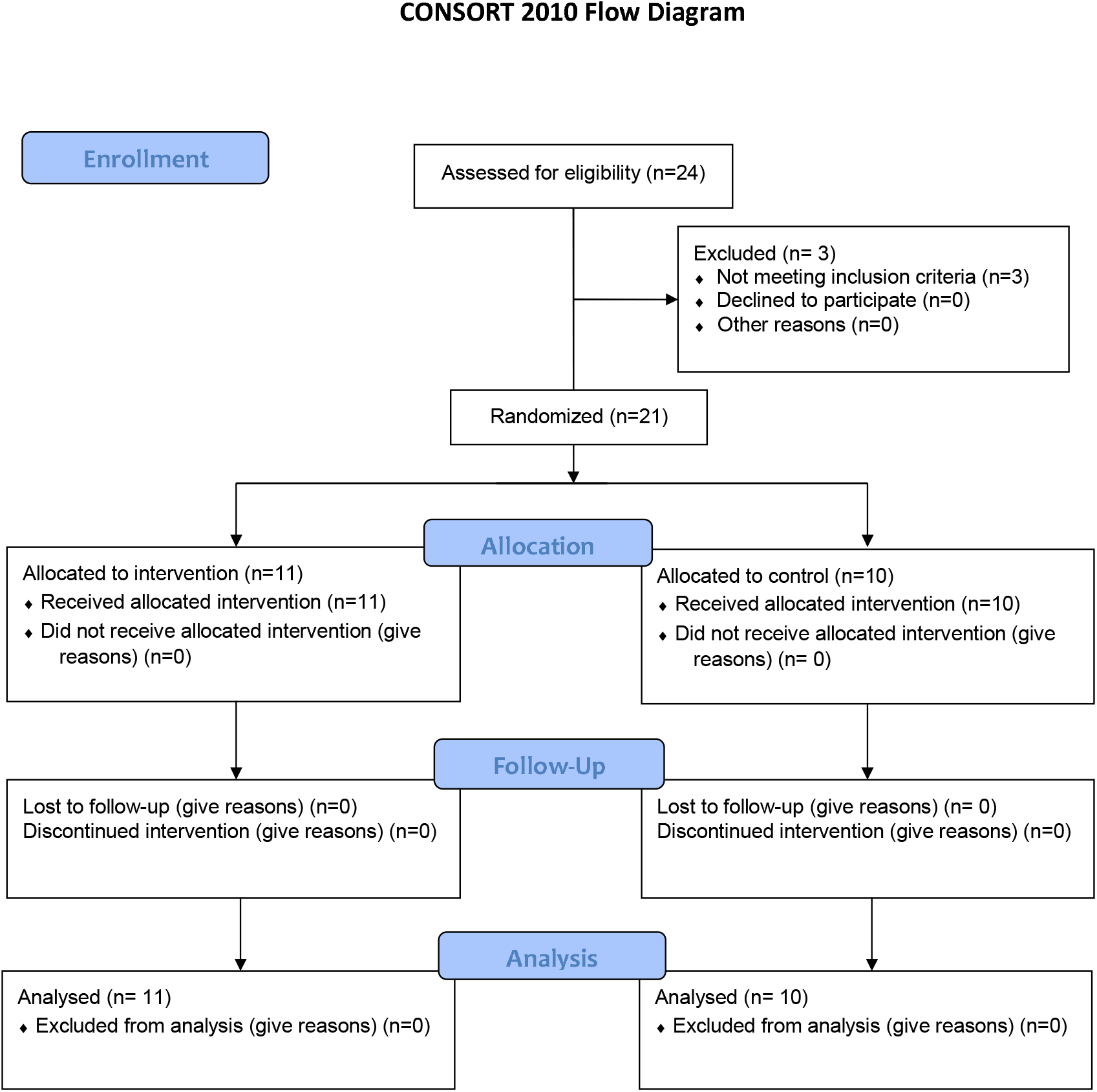
CONSORT flow diagram of enrollment, allocation, follow-up, and analysis.

The study was terminated early due to safety concerns identified in the first 20 participants upon interim analysis. The DMC recommended discontinuation, and the trial steering committee agreed accordingly. We analyzed the remaining outcomes using aggregated data from all 21 participants included in the study. We recorded no participant withdrawals, no loss to follow-up, or missing data in either the final or interim analysis. All participants completed follow-up except for four in each group who died. Baseline characteristics were balanced between groups (see Table 1). In nine out of 17 cases (52.9%) with a Graeb score of 5 or above, bilateral ventricular catheter treatment was administered due to Monro block. The number of patients with severe IVH was equally distributed between the two groups (mean Graeb score 8.7 versus 9.3, p=0.72).

### Safety outcomes

Based on an interim analysis of the first 20 participants (10 treated with IRRAflow^®^; 10 with EVD), seven out of ten IRRAflow^®^ patients (70%) experienced at least one SAE compared with two out of ten (20%) in the EVD group (p=0.024). Therefore, the study was terminated early. Please see Supplementary Material 6-9 for root cause analysis, complete DMC safety report, frequencies, and AU causality assessments. Table 2 presents safety outcomes for the final analysis population. The risk of experiencing at least one SAE was significantly higher in the IRRAflow^®^ group (63.6%) than in the EVD group (20%), with an absolute risk difference of 0.43 (95% CI 0.059-0.813, p=0.044). Similarly, the SAE incidence was significantly higher with IRRAflow^®^ (incidence rate ratio = 6.0, 95% CI 1.38-26.11, p=0.012).

**Table 2.** Safety and efficacy outcomes.

|  | Intervention | Control | Comparison | p-value |
| --- | --- | --- | --- | --- |
| **Safety outcomes** |  |  |  |  |
| Risk of $\geq 1$ SAE | 63.6%<br>(95%CI 42.0-98.4) | 20%<br>(95%CI 4.8-44.8) | Risk difference<br>0.43, 95%CI (0.059-0.813) | **0.044\*** |
| No. of SAEs | 17 | 2 | Incidence rate ratio<br>6.0 95%CI (1.38-26.11) | **0.012\*** |
| Catheter related infections | 18.2%<br>(95%CI 5.2-63.7) | 10%<br>(95%CI 1.6-64.2) | Risk ratio<br>1.82 95%CI (0.19-17.1) | 0.601 |
| 30-day mortality rate | 27% | 40% | Risk difference<br>12.7% 95%CI (-52.9-27.5) | 0.535 |
| **Efficacy outcomes** |  |  |  |  |
| Catheter occlusion, all catheters | 37.5% | 6.6% | Hazard ratio<br>4.4 95%CI (0.6-31.2) | 0.141<sup>1**</sup> |
| Catheter occlusion, primary catheter | 36.4% | 10% | Hazard ratio<br>3.2 95%CI (0.35-28.7) | 0.303 |
| Procedure time (range) | 53.5 min<br>(36-116 min) | 12 min<br>(4-32 min) | Ratio<br>4.5 95%CI (2.6-10.4) | **0.0001\*** |
| Clearance of blood, clot half life | 3.9 days<br>95%CI (1.25-6.57) | 5.4 days<br>95%CI (-0.17-10-9) | Difference<br>1.5 95%CI (-4.69-7.6) | 0.643 |
| Shunt dependency | 18.2%<br>95%CI (5.2-63.7) | 10%<br>95%CI (1.6-64.2) | Risk ratio<br>1.82 95%CI (0.19-17.1) | 0.601 |
| Length of stay, ICU (mean) | 17.5 days<br>95%CI (11.1-23.8) | 10.9 days<br>95%CI (0.2-22.0) | Difference<br>6.55 days (5.6-18.6) | 0.266 |
| 90 days mRs $\leq 3$ | 90.9%<br>95%CI (75.4-109.6) | 90.0%<br>95%CI (73.2-110.7) | Risk ratio<br>1.01 95CI (0.76-1.33) | 0.944 |
| Duration of catheter treatment (mean) | 11.5 days<br>(SD 6.6) | 11 days<br>(SD 8.6) | Difference<br>0.5 95%CI (-6.5-7.6) | 0.873 |

\*significant result (5% significance level), \*\* significant result (20% significance level), <sup>1</sup> p-value based on Hazard ratio (not the absolute risks of occlusion)
CI = Confidence interval
ICU = Intensive care unit
mRs = modified Rankin Scale
SAE = Severe adverse events

### Efficacy outcomes

The IRRAflow^®^ group had a significantly higher catheter occlusion rate (primary outcome) than the EVD group (37.5% versus 6.6%) (hazard ratio 4.4 (95%CI 0.6-31.2, p=0.141, meeting the prespecified 0.2 alpha level) (Figures 3a & b). Among intervention group occlusions, 60% received a contralateral EVD and 40% were treated with tPA for clot resolution or had catheter replacement. In the control group, the single patient (10%) experiencing occlusion had catheter replacement.

**Figure 3.**
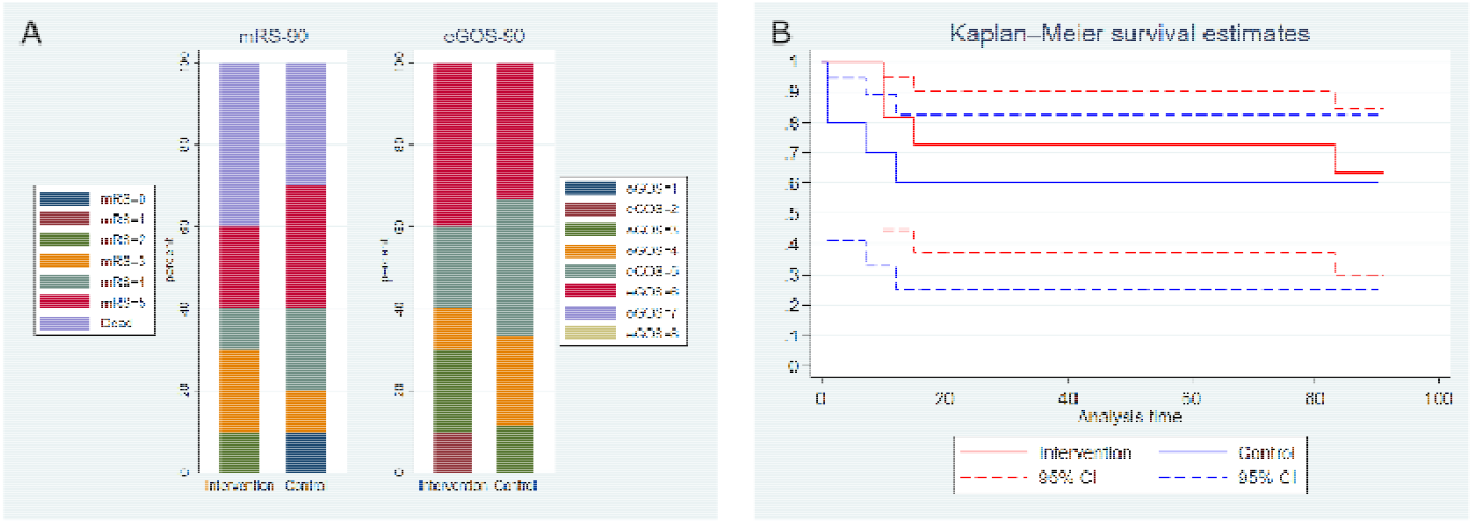
Functional outcome and survival. A. Bar plot of the extended Glasgow Outcome Scale score and modified Rankin Scale score at 90 days for the two groups (risk ratio for good functional outcome mRS <3 = 1.01, 95%CI (0.76-1.33), p=0.944). B. Kaplan-Meier curves of survival in the two groups. Log-rank test for equality of survival comparing the two survival curves showed no difference in survival between the two groups (p=0.672 and 30-day mortality rate: 27% versus 40, risk difference 12.7% 95%CI (−52.9-27.5), p=0.535).

The IRRAflow^®^ group had a significantly longer median procedure time for catheter placement than the EVD group (53.5 min versus 12 min, p=0.0001). ICU admission tended to be longer with IRRAflow^®^ but the difference was statistically insignificant (mean length of ICU stay 17.5 days (95%CI 11.1-23.8) versus 10.9 days (95%CI 0.02-22), p=0.266). Hematoma clearance rate did not differ significantly between the IRRAflow^®^ group and the control group (clot half-life = 3.9 days (95%CI 1.25-6.57) versus 5.4 days (95%CI -0.17-10.9), p=0.643).

The mean irrigation volume in the intervention group was 1,093 ml/day (range 0-4,166). Additional secondary outcomes are provided in Table 2 and Figure 4a&b.

## Discussion

IVH is a severe condition causing significant neurological complications, including brain damage, disability, and death. Although introduction of IVF treatment nearly two decades ago has facilitated clot removal and improved patient outcome, mortality rates remain in the 20-50% range, and more than half of survivors live to achieve poor functional outcome (mRS > 3)^2^. No significant recent treatment advances have been made to improve patient outcomes, and new technologies in the field are urgently needed. Various minimally invasive techniques have been suggested and are recommended by AHA guidelines as they have shown reductions in mortality rates^8^.

In this randomized, we conducted a controlled evaluation of the safety and efficacy of the IRRAflow^®^ technology (IRRAS AB) for active wash-out of IVH compared with standard-practice passive EVD. IRRAflow^®^ builds on the novel and innovative principle of controlled intraventricular perfusion, providing a potential and critical leap forward in hemorrhagic stroke treatment. The method proposedly facilitates IVH clearance and reduces the risks of catheter clotting and drain-related CNS infections, thereby addressing key issues of IVH treatment. Despite its potential, the study was terminated early due to safety concerns with IRRAflow^®^ as recommended by an independent DMC. Our findings revealed a significant six-fold increase in SAE incidence with IRRAflow^®^ compared with EVD. Furthermore, contrary to the primary outcome hypothesis, a significant 4.4-fold increase was recorded in the risk of catheter occlusion with IRRAflow^®^.

An extensive individual root cause analysis revealed that the majority of SAEs were causally related to IRRAflow^®^ device features, such as 1) small inner drain line lumen, 2) lack of antibiotic catheter coating, 3) repeated exchange of saline supply for irrigation, 4) insufficient catheter fixation method, 5) incompatibility with neuronavigation systems, 6) an excessively negative pressure gradient during the aspiration phase, 7) and error-prone ICP measurements and supervisory control. Actions should be taken to mitigate these caveats and potential treatment hazards in future versions of the technology^1,9-12^. Furthermore, precise guidelines for safe, feasible, and effective parameter settings of IRRAflow^®^ are needed to streamline and simplify treatment.

Except for the safety outcomes, most endpoints in the study did not reach statistical significance, as expected due to early termination and reduced statistical power. Mortality, functional outcome, shunt-dependency, and hematoma clearance showed no significant effects. However, a significant 4.5-fold increase was recorded in surgery time for IRRAflow^®^ despite proper training and study personnel certification. This underscores the need to simplify the surgical procedure and device setup as CSF drainage is an acute and potentially life-saving treatment for IVH. Preliminary endpoint assessments may be used for future reference and study designs.

Although our findings oppose the use of IRRAflow^®^ for IVH under the given circumstances, the principle of intraventricular lavage may still harbor promise for improved IVH clearance and patient outcome. Faster clearance of blood has been shown to significantly reduce mortality rates, increase functional outcome, and decrease catheter occlusion rates^1^. A combination of IVF and IRRAflow^®^ may potentially increase blood clearance compared with irrigation alone or IVF alone.

Currently, only one published study, DRIFT, has evaluated IRRAflow^®^ treatment of IVH patients^4^. The study is a case series of four patients treated with IRRAflow^®^ in combination with IVF. The authors found clear radiographic and clinical superiority of IRRAflow^®^ and observed no complications compared with standard EVD4. Hence, the combination of IRRAflow^®^ and IVF is potentially safe and feasible, which is further investigated in the ongoing ARCH study (NCT05118997). In this study, IRRAflow^®^ plus continuous IVF infusion and IRRAflow^®^ plus manually administered IVF are being tested against standard EVD + IVF. This and other studies will shed a broader light on the safety, feasibility, and efficacy of IRRAflow^®^ under various conditions.

## Study limitations

First, the nature of IRRAflow^®^ surgery and treatment made blinding of treating physicians and incorporation of a sham device impossible. Second, 87.7% of the final analysis population had a Graeb score of 5 or above corresponding to severe IVH, which is associated with a poorer outcome than Graeb scores 1-4^13^. This potentially compromises generalizability of the results to less severe IVH conditions. Third, the study enrolled patients with primary and secondary IVH, including SAH, collectively representing a relatively inhomogeneous population. This was not a problem for assessing general safety, feasibility, and efficacy surrogate markers, such as IVH clearance and catheter obstruction for IRRAflow^®^. However, future studies on clinical efficacy endpoints should be designed for homogeneous patient subcategories. Finally, participants with Monro block were treated with standard EVD in the contralateral ventricle, regardless of randomization. This may potentially disturb intent-to-treat results, although most AEs were found to be associated directly with IRRAflow^®^ features. However, future studies may consider investigating bilateral use of IRRAflow^®^ in case of Monro block.

## Conclusion

We found that the current IRRAflow^®^ technology was encumbered with serious caveats resulting in a significantly increased SAE risk under the investigated conditions. However, we believe that the principle of controlled intraventricular lavage potentially holds promise for improvement of IVH treatment.

## Supporting information

All supplemenatry

## Data Availability

All data produced in the present work are contained in the manuscript

## Acknowledgements

We sincerely thank the entire neuro Intensica care unit (ICU) staff for competent and professional implementation and handling of trial procedures, particularly nurses Fie Jappe and Kirsten Loch from Aarhus University Hospital, and nurses Maria Halle and Helle Dixen Hansen from Odense University Hospital. Furthermore, we take this opportunity to express our gratitude to all the nurses from the Department of Neurosurgery, particularly Ane Kirkegaard, and to IRRAS AB for competent staff training and education.

## Other information

### Registration

ClicalTrials.gov identifier: NCT05204849, registered 15 December 2021, updated 24 January 2022

### Protocol

The published full protocol can be found https://doi.org/10.1186/s13063-022-07043-9 or in Supplementary Material 1

