## Supplementary material for "Safety, harm, and efficacy of IRRAflow^®^ versus external ventricular drainage for intraventricular hemorrhage: A randomized clinical trial": All supplemenatry: Supplementart 7 Root cause analysis Medrxiv.docx

**Supplementary material 7. Root cause analysis of all SAEs**

This root cause analysis (RCA) is based on the “five why” RCA method and based on following steps: 1) problem definition, including a description of the event and formation of a problem statement. 2) Gathering of data, including an investigation of the incident and collection information regarding the event. 3) Data analysis with identification of possible causal factors for the event. 4) Identification of root causes of the causal factors using the “five why” method. 5) Recommendation for corrective actions.

| Problem definition  Description of the event and formation of a problem statement. | Data gathering  Incident investigation: Collection of information regarding the event. | Causal factors  Causal factors causing the event | Root causes  The five-way analysis of root causes | Recommendations  Preventive action to prevent event from happening again |
| --- | --- | --- | --- | --- |
| **INTERVENTION GROUP** |  |  |  |  |

**Patient ID 1**

Enrolment of January 2022	No SAEs observed

- ICH with break through
- Graeb score 6
- Unilateral catheter treatment

| **Patient ID 2**  Enrolment January 2022   - ICH with break through - Graeb score 8 - Bilateral catheter treatment - Both EVDs placed during same procedure after admission - Left lateral ventricle: IRRAflow catheter - Right lateral ventricle: standard external ventricular drain (EVD) | | | | |
| --- | --- | --- | --- | --- |
| Problem 1)  Event: SAE grade 3, displacement - irrigation into parenchyma  IRRAflow control unit showed negative ICP values (-20 mmHg) compared to control Raumedic ICP monitor (ICP 19-23 mmHg).  Hence, the IRRAflow was inconstantly working, irrigation intermittently functioning and drainage intermittent functioning.  Meanwhile the EVD on contralateral site was closed for potential removal reopened due to malfunctioning IRRAflow and ICP above 20 mmHg on raumedic device.  **Problem statement**  Malfunctioning IRRAflow: Incorrect IRRAflow control unit ICP values causing increase in ICP due to only intermittent drainage. | Due to ICP increase and malfunctioning IRRAflow a head CT scan was performed. CTC showed displacement of the catheter just lateral to the ventral horn of the left lateral ventricle.  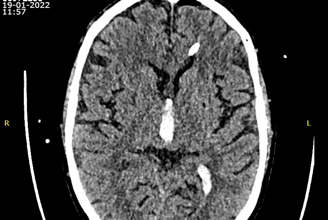  Hence, drainage was only possible when the side-holes of the catheter tip were in contact to cerebrospinal fluid (CSF). Further, due to the displacement Irrigation had happened into the brain parenchyma.  EVD on contralateral side was reopened for ICP control. | Catheter displacement  IRRAflow device failure | **Catheter displacement**  #1 Why was the IRRAflow malfunctioning?  The catheter tip had displaced into the brain parenchyma.  #2 Why was the catheter tip displaced into the brain parenchyma?  The IRRAflow catheter is tunneled and fixed by a fixation cap and with sutures.  #3 Why did that specific fixation method cause displacement?  The fixation cap only fixates the catheter very loosely. Sutures is not allowed to be too tight as catheter is then at risk of occlusion.  #4 Why did the displacement cause malfunctioning?  The side-holes of the IRRAflow catheter were displaced into the parenchyma and not surrounded by CSF.  #5 Why is the IRRAflow catheter tip dependent on being surrounded by liquid?  The IRRAflow ICP monitor is based on hydrostatic pressures requiring liquid around the side-holes of the catheter tip.  **IRRAflow device failure**  #1 Why did the IRRAflow show incorrect ICP values?  The side-holes of the catheter must be surrounded by liquid to evaluate the ICP  #2 Why do the side-holes have to be surrounded by liquid?  The IRRAflow ICP monitor is based on hydrostatic pressures requiring liquid around the side-holes of the catheter tip.  #3 Why was there no liquid around the catheter tip?  The catheter tip had displaced into the parenchyma  #4 Why did the catheter tip displace into the parenchyma?  The IRRAflow catheter is tunneled and fixed by a fixation cap and with sutures.  #5 Why is the catheter not better fixated?  The catheter is incompatible to a bolt due to missing metal enforcement. | **Most likely cause of event**  Catheter displacement due to fixation method  **Recommendation**  The catheter should be placed on a bolt which would decrease the risks of displacements. |
| Problem 2)  Event: SAE grade 3, displacement – irrigation into parenchyma  IRRAflow completely incapacitated drainage. No drainage, intermitted irrigation and invalid ICP values.  **Problem statement**  Malfunctioning IRRAflow: Irrigation into brain parenchyma, no drainage. | Control head CT revealing further displacement of catheter and at this point no connection the lateral ventricle explaining the drainage stop.  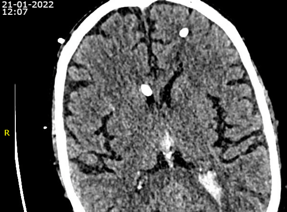  Irrigation had happened into the brain parenchyma.  EVD on contralateral side ensuring ICP control. | Catheter displacement  IRRAflow device failure | **Catheter displacement**  #1 Why was the IRRAflow malfunctioning?  The catheter tip had displaced into the brain parenchyma.  #2 Why was the catheter tip displaced into the brain parenchyma?  The IRRAflow catheter is tunneled and fixed by a fixation cap and with sutures.  #3 Why did that specific fixation method cause displacement?  The fixation cap only fixates the catheter very loosely. Sutures cannot be too tight as catheter is in risk of occlusion.  #4 Why did the displacement cause malfunctioning?  The side-holes of the IRRAflow catheter were displaced into the parenchyma and not surrounded by cerebrospinal fluid (CSF).  #5 Why is the tip of the IRRAflow catheter dependent on being surrounded by liquid?  The IRRAflow ICP monitor is based on hydrostatic pressures requiring liquid around the side-holes of the catheter tip.  **IRRAflow device failure**  #1 Why did the IRRAflow show incorrect ICP values?  The side-holes of the catheter must be surrounded by liquid to evaluate the ICP  #2 Why do the side-holes have to be surrounded by liquid?  The IRRAflow ICP monitor is based on hydrostatic pressures requiring liquid around the side-holes of the catheter tip.  #3 Why was there no liquid around the catheter tip?  The catheter tip had displaced into the parenchyma  #4 Why did the catheter tip displace into the parenchyma?  The IRRAflow catheter is tunneled and fixed by a fixation cap and with sutures.  #5 Why is the catheter not better fixated?  The catheter is incompatible to a bolt due to missing metal enforcement. | **Most likely cause of event**  Catheter displacement due to fixation method  **Recommendation**  The catheter should be placed on a bolt which would decrease the risks of displacements. |
| Problem 3)  Event: SAE grade 3, ventriculitis  Persistent elevated temperature above 38 degrees Celsius with no response to antibiotics. The patient was treated with a craniotomy for hematoma evacuation The patient was treated with bilateral catheters (IRRAflow and EVD). The IRRAflow was pushed a few cm deeper due to a small displacement. The patient was priorly treated for pneumonia four days before event.  **Problem statement**  Fever with no infectious focus | Due to persistent body temperature and no response to antibiotics a sample of CSF was analysed and found positive for bacteria (staphylococci epidermidis).  The patient was treated with intra thecal vancomycin 20 mg x 1 pr. day. | Catheter related infection:   - IRRAflow catheter related - Contralateral EVD related   Hematogenous spread or postoperative infection after craniotomy | **Infection related to catheter:**  #1 Why is the infection related to catheter treatment?  Foreign objects are at risks of biofilm coating and are very bacteriophages.  #2 Why is the EVD at risk of causing infection? The EVD is silver impregnated which is shown to be less antibacterial compared to antibiotic coating. However, the EVD was placed on a bolt which reduce risks of infection in comparison to tunneled catheters.  #3 Why is the IRRAflow at risk of causing the infection?  The IRRAflow catheter is uncoated by neither antibiotics nor silver and placed by reverse tunneling.  #4 Why does tunneling increase risks of infection?  Tunneling is shown to increase risks of infection compared to bolted EVDs. This is due to the canal along the catheter, risks of CSF leakage and small slidings of the catheter.  #5 Why is the IRRAflow the potential cause of the infection even though the IRRAflow catheter was removed before the finding of the infection?  The IRRAflow was removed 21^st^ of January. The positive sample was taken 23^rd^ of January. However, growth of bacteria take days and causality to IRRAflow is therefore still highly relevant^1^.  **Hematogenous spread or postoperative infection after craniotomy:**    #1 Why could the infection origin from hematogenous spread?  The patient had a positive tracheal sample  #2 Why was hematogenous spread not primary suspicion of infectious focus?  The bacteria found here were Klebsiella Pneumoniae whereas the bacteria causing the ventriculitis were staphylococci epidermidis.  #3 Why was an alternative primary infection not a possibility?  An alternative primary focus was not found or suspected.  #4 Why could the infection be related to the craniotomy?  Low virulent infections could potentially have caused the positive CSF sample 8 days postoperatively.  #5 Why was the craniotomy less likely have caused the infection:  The scar was healing nicely and no signs of infection in relation to deeper layers of the scar or craniotomy was observed on head CT scans. | **Most likely cause of event**  IRRAflow catheter treatment due to:   - Reverse tunneling of catheter - Uncoated catheter - Fixation: Catheter was pushed deeper and later displaced again   **Recommendation:**  The IRRAflow catheter should be placed on a bolt and be antibiotic coated. |
| Problem 4)  Event: SAE grade 5, died  **Problem statement**  Patient passed away | The patient was transferred to a rehabilitation facility. During rehabilitation no improvement in consciousness achieved. The patient was very bothered by the tracheal tube which was removed accompanied by an aspiration pneumonia. Active treatment was ended, and the patient died April 2022 due to poor respiratory conditions. | Poor outcome after severe hemorrhage | **Poor outcome after sever hemorrhage**  #1 Why did the patient die  Due to aspiration pneumonia  #2 Why did the patient experience an aspiration pneumonia  Due to extubation  #3 Why was the patient extubated  Due to bothersome symptom from the tube  #4 Why was bothersome symptoms indication of tube removal  Due to poor prognosis  #5 Why was the prognosis poor?  Low level of conscious and no improvement during rehabilitation. | **Recommendations:**  None |

| **Patient ID 6**  Enrolment March 2022   - SAH with break through - Graeb score 3 - Unilateral catheter treatment | | | | |
| --- | --- | --- | --- | --- |
| Problem 1)  Event: SAE grade 5, died  **Problem statement**  Patient passed away | Severe SAH and poor prognosis from the beginning. After 3 days without sedation and a negative EEG all active treatment was ended, and the patient died expectedly. | Poor prognosis after severe hemorrhage | **Poor prognosis after severe hemorrhage**  #1 Why did the patient die?  Due to end of active treatment  #2 Why was the active treatment ended?  Due to very poor prognosis  #3 Why was the prognosis poor?  The patient had experienced a very severe SAH and was observed for 3 days without sedation with no sign of awakening.  #4 Why was there no sign of awakening? Vegetative state due to severe brain damage. EEG was found to be normal.  #5 Why was the EEG performed?  An electro encephalogram (EEG) was performed to ensure that the vegetative state was not due to seizures. | **Recommendations:**  None |

| **Patient ID 8**  Enrollment March 2022   - SAH with break through - Graeb score 11 - Bilateral catheter treatment - Right lateral ventricle: IRRAflow catheter - Left lateral ventricle: EVD | | | | |
| --- | --- | --- | --- | --- |
| Problem 1)  Event: SAE grade 3, catheter replacement  After catheter placement in the operating room (OR) the patient was transferred to endovascular coil treatment of an aneurism. After the procedure the catheter was displaced and found on the patients pillow. The patient was transferred back to the OR for a new IRRAflow catheter placement.  **Problem statement**  Catheter on pillow outside of the patient. Catheter was replaced during a new surgical procedure | During the coil procedure the patient was moved from one bed to another and further moved back and forth during the scans which was performed during the coil procedure. After the coil procedure was finished the catheter was found beside the patient on the pillow. | Catheter fixation method  Length of IRRAflow tube set | **Catheter fixation method**  #1 Why could the fixation method cause the displacement?  The IRRAflow is placed by tunneling and fixed using a fixation cap and sutures.  #2 Why are tunneled, suture fixated catheter more likely of displacing?  Catheter fixed by sutures are more likely to displace compared to bolted EVD as they are not as securely fastened as bolted catheters.  #3 Why is bolted EVDs more securely fastened?  The bolt is screwed through the cranium and the bolt is screwed until the catheter is securely fastened.  #4 Why is the IRRAflow not placed on a bolt?  The IRRAflow catheter lacs a metal enforcement enabling secure fastening of the bolt around the catheter.  #5 Why is the metal enforcement necessary?  The metal enforcement prevents the catheter lumen form constriction and there by occlusion.  **Length of IRRAflow tube set**  #1 Why could movement before and during coil procedure cause the event?  The movement could potentially cause an unobserved pull in the tubes and thereby in the catheter causing the catheter to displace and be pulled out of the patient.  #2 Why could the pull cause the displacement?  The tubes of the IRRAflow are relatively short and the IRRAflow control unit is therefore very importantly placed close to the patient’s head.  #3 Why could a pull cause the displacement?  The IRRAflow is placed by tunneling and fixed using a fixation cap and sutures. Catheter fixed by sutures are more likely to displace compared to bolted EVD.  #4 Why is bolted EVDs more securely fastened?  The bolt is screwed through the cranium and the bolt is screwed until the catheter is securely fastened.  #5 Why is the IRRAflow not placed on a bolt?  The IRRAflow catheter lacs a metal enforcement enabling secure fastening of the bolt around the catheter without occluding the catheter lumens. | **Most likely cause:**  A combination of the catheter fixation method and short tube sets of the IRRAflow.  **Recommendations:**   - The IRRAflow catheter should be compatible to bolted placement. - The tube set of the IRRAflow should be longer. |
| Problem 2)  Event: SAE grade 3, catheter replacement  The patient was treated with bilateral catheters. The EVD occluded and was replaced. Further alteplase administrated to lower risks of new occlusions.  **Problem statement**  Catheter occlusion | The left standard EVD was observed to be occluded. No drainage was observed and the EVD has stopped oscillating. The IRRAflow on the right side was stable and well-functioning (reliable ICP and drainage).  Due to occlusion the patient was transferred to the OR and the left standard EVD was replaced using the same bolt and parenchymal canal. Tissue plasminogen activator (tPA) was administrated to avoid further occlusions. | Blood clots | **Blood clots**  #1 Why was the catheter replaced?  Due to no drainage.  #2 Why was the catheter malfunctioning?  Due to occlusion  #3 Why was the catheter occluded?  The patient had suffered a massive IVH with blood in all parts of the ventricles. Under these circumstances occlusion is very likely to happen.  #4 Why was tPA administrated?  To reduce risks of occlusion of the new catheter  #5 Why does tPA reduce risks of catheter occlusions?  tPA converts plasminogen to the active form plasmin facilitates clot resolution^2^. tPA has been found to significantly decrease risks of occlusions in IVH patients. | **Most likely cause:**  Occlusion due blood clots  **Recommendations:**  Early tPA administration to avoid catheter occlusions. |
| Problem 3)  Event: SAE grade 3, ventriculitis  Elevated temperature at 38.8 degrees Celsius.  The patient was treated with bilateral catheters.   - IRRAflow right side - EVD left side   **Problem statement**  Fewer and no infectious focus | Due to elevated body temperature a sample of CSF was analysed and found positive for bacteria (coagulase negative staphylococci).  The patient was treated with IT vancomycin 20 mg x 1 pr. day. | Catheter related infection:   - IRRAflow catheter related - Contralateral EVD related   Hematogenous spread | **Infection related to catheter treatment:**  #1 Why was the infection related to catheter treatment?  Foreign objects are at risks of biofilm coating and are very bacteriophages.  #2 Why was the EVD at risk of causing infection? The EVD is silver impregnated which is shown to be less antibacterial compared to antibiotic coating. However, the EVDs are placed on a bolt which reduce risks of infection in comparison to tunneled catheters.  #3 Why was the IRRAflow at risk of causing the infection?  The IRRAflow catheter is an uncoated catheter with neither antibiotic nor silver impregnation and placed by reverse tunneling.  #4 Why was reverse tunneling a risk factor for infection?  Tunneling of catheters are shown to increase risks of infection compared to bolted catheters. The canal along the catheter, the risks of CSF leakage and the small slidings of the catheter are thought to increase the risks.  #5 Why was the IRRAflow the potential cause of the infection even though the IRRAflow catheter was removed before the finding of the infection?  The IRRAflow was removed 11^st^ of April. The positive sample was taken 14^th^ of April. However, growth of bacteria take days and causality to IRRAflow is therefore still highly relevant^3^.  **Hematogenous spread:**  #1 Why could the infection origin from hematogenous spread?  The patient had a positive tracheal sample, however bacteria found here were Haemophiles Influenza.  #2 Why was an alternative primary infection not a possibility?  An alternative primary focus was not found or suspected. | **Most likely cause of event**  IRRAflow catheter treatment due to:   - Reverse tunneling of catheter - Uncoated   **Recommendation:**  IRRAflow catheter should be placed on a bolt and be antibiotic coated. |

| **Patient ID 9**  Enrolment April 2022   - SAH with break through - Graeb score 3 - Unilateral catheter treatment | | | | |
| --- | --- | --- | --- | --- |
| Problem 1)  Event: SAE grade 3, ventriculitis  Elevated temperature 39.0 degrees Celsius. Primary suspicion of focus was CNS due to fluctuating GCS scores.  **Problem statement**  Fewer with primary CNS focus suspicion | CSF samples were taken from the patient due to elevated temperature and fluctuating GCS. The samples were found positive for staphylococcus aureus.    Due to the positive CSF culture the patient is transferred to the OR and the IRRAflow catheter was replaced to a tunnelled Silverline catheter through the old cicatrice from the IRRAflow placement. Patient was treated with IV Cefuroxime 3000 mg x 3 pr. day and IT vancomycin 20 mg x 1 pr day. | Catheter related infection | **Infection related to catheter treatment:**  #1 Why was the infection related to catheter treatment?  Foreign objects are at risks of biofilm coating and are very bacteriophages.  #2 Why was the IRRAflow at risk of causing the infection?  The IRRAflow catheter is an uncoated catheter with neither antibiotic nor silver impregnation and placed by reverse tunneling.  #3 Why are uncoated catheters related to higher risks of infection?  Patients with catheter coated with antibiotic agents are significant less likely to be infected due to the antibacterial impregnation of the catheter.  #4 Why was reverse tunneling a risk factor for infection?  Tunneling of catheters are shown to increase risks of infection compared to bolted catheters. The canal along the catheter, the risks of CSF leakage and the small slidings of the catheter are thought to increase the risks.  #5 Why was no other focus considered?  The patient had tracheal and urine samples evaluated with no sign of infection. Furter chest x-ray was negative. | **Most likely cause of event**  IRRAflow catheter treatment due to:   - Reverse tunneling of catheter - Uncoated catheter   **Recommendation:**  IRRAflow catheter should be placed on a bolt and be antibiotic coated. |
| Problem 2)  Event: SAE grade 3, catheter replacement  The catheter had been closed to evaluate whether the catheter could be removed without signs of hydrocephalus. Due to decrease in consciousness the catheter was reopened. However, the catheter has meanwhile occluded, and no drainage or oscillation was observed.  **Problem statement**  Catheter occlusion | IRRAflow catheter was replaced with a tunnelled Silverline catheter due to ventriculitis.  The tunnelled Silverline catheter showed no signs of oscillation nor drainage. The patient was descended in GCS and clinically in need of CSF drainage. The patient was transferred to the OR and had a new bolted catheter placed on the contralateral side. | Catheter occlusion | **Catheter occlusion**  #1 Why was the catheter replaced?  Due to malfunctioning with no CSF drainage nor oscillation.  #2 Why was the catheter malfunctioning?  Due to occlusion of the catheter  #3 Why was the catheter occluded?  The catheter could potentially be occluded by blood clots. However, the patient was 19 days after ictus and composed blood clots were no longer found in the ventricles.  #4 Why did the patient then experience an occlusion?  Most likely due to brain debris.  #5 Why did brain debris cause occlusion?  Brain debris are found to cause occlusion of catheters and seemed to be the reason for the occlusion. | **Most likely cause of event:**  Occlusion due to debris of the brain  **Recommendation:**  The IRRAflow catheter should have a larger inner diameter. Catheters should be left as short as possible. For patients with persistent need of CSF drainage a VP-shunt should be placed as soon as clinically possible. |

| **Patient ID 11**  Enrolment May 2022   - ICH with break through - Graeb score 7 - Bilateral catheter treatment - Right lateral ventricle: IRRAflow catheter - Left lateral ventricle: EVD | | | | |
| --- | --- | --- | --- | --- |
| Problem 1)  Event: SAE grade 3, contralateral catheter placement  The patient was randomised to IRRAflow treatment, and the catheter was placed just before midnight on the right side. The patient was afterwards transferred to a head CT to confirm catheter placement with tip in the ventricles. CT confirmed correct placement. After the CT scan, the patient was transferred to the intensive care unit and drainage and irrigation was initiated. However, no drainage of CSF was observed even though the ICP was increased to 25-30 mmHg.  **Problem statement**  Catheter occlusion, EVD placed contralaterally. | Just after IRRAflow placement the IRRAflow control unit was malfunctioning with no observed drainage. The head CT revealed that the tip of the catheter lied in hematoma and thus the catheter was occluded. ICP increase to 25-30 mmHg. Further incorrect ICP values were intermittently observed on the IRRAflow control unit. The patient was transferred back to the OR for EVD placement on contralateral side for ICP control.  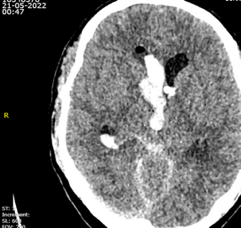 | Occlusion due to blood clots  Device failure  Diameter of catheter lumen  Aspiration method of the IRRAflow | **Occlusion due to blood clots**  #1 Why was an additional EVD placed contralaterally?  Due to IRRAflow malfunctioning.  #2 Why was the IRRAflow malfunctioning?  The catheter tip was placed in hematoma.  #3 Why did the catheter localization in hematoma cause drainage stop  The catheter was occluded.  #4 Why did the catheter localization cause incorrect ICP values?  The side-holes of the IRRAflow catheter were placed in hematoma and not surrounded by CSF.  #5 Why was the tip of the IRRAflow catheter dependent on being surrounded by liquid?  The IRRAflow ICP monitor is based on hydrostatic pressures requiring liquid around the side-holes of the catheter tip.  **IRRAflow device failure**  #1 Why did the IRRAflow show incorrect ICP values?  The side-holes of the catheter must be surrounded by liquid to evaluate the ICP  #2 Why do the side-holes have to be surrounded by liquid?  The IRRAflow ICP monitor is based on hydrostatic pressures requiring liquid around the side-holes of the catheter tip.  #3 Why was there no liquid around the catheter tip?  The catheter tip was surrounded by hematoma  **Diameter of catheter lumen**  #1 Why was the risks of occlusion related to the diameter of the IRRAflow catheter?  The inner diameter of the IRRAflow drainage line is 1.5 mm whereas the inner diameter of a standard 10 fr Silverline catheter is 1.9 mm.  #2 Why is the IRRAflow drainage line inner diameter smaller than the inner diameter of an EVD?  The IRRAflow catheter is a dual lumen catheter with an irrigation line and drainage line.  #3 Why is the risks of occlusion potentially higher in the IRRAflow catheter compared to a standard EVD?  Small lumen catheter is at significantly higher risks of occlusions than large diameter catheters  #4 Why does the irrigation of the IRRAflow no avoid occlusions?  The catheter is divided in two lumens and only the tip is a combined lumen which is held open by irrigation. The inner part of the catheter is separated, and the drainage line is therefore not held open by the irrigation.  **Aspiration method of the IRRAflow**  #1 Why could the occlusion be due to the aspiration method of the IRRAflow?  The IRRAflow performs suction instead of passive drainage as the drainage bag is placed from 0 to -103 cm below the patient causing a negative pressure gradient with accompanied suction.  #2 Why is suction needed when using the IRRAflow device  It is thought that when irrigation a volume of saline into the ventricles it should be followed by a suction mechanism to make sure of sufficient drainage.  #3 Why could suction cause occlusion  Suction can cause collapse of the ependyma around the catheter tip or by sucking the hematoma through the holes in the catheter tip. | **Most likely cause of event:**  Occlusion due to location of catheter tip in hematoma.  **Recommendation:**  The inner diameter of the drainage line in the IRRAflow catheter would favorably be increased. Catheter should be able to function inside the hematoma and maybe be accompanied by tPA administration. |
| Problem 2)  Event: SAE grade 3, tPA administration to avoid severe hydrocephalus  The IRRAflow showed incorrect ICP values from -10 to 100 mmHg. No drainage was observed. The IRRAflow control unit is designed to stop treatment when ICP values exceeds device alarm limits of -5 to 35 mmHg.  **Problem statement**  Catheter occlusion, tPA administrated | ICP values measured in IRRAflow device showed fluctuating values between -10 to 100 and no drainage was observed. Head CT showed that the catheter tip was placed in hematoma. tPA was administrated to resolve the clot and occlusion of the catheter to avoid severe hydrocephalus. | Occlusion due to blood clots  Device failure  Diameter of catheter lumen  Aspiration method of the IRRAflow | **Occlusion due to blood clots**  #1 Why was tPA administrated?  Due to IRRAflow malfunctioning.  #2 Why was the IRRAflow malfunctioning?  The catheter tip was placed in hematoma.  #3 Why did the catheter localization in hematoma cause drainage stop  The catheter was occluded.  #4 Why did the catheter localization cause incorrect ICP values?  The side-holes of the IRRAflow catheter were placed in hematoma and not surrounded by CSF.  #5 Why was the tip of the IRRAflow catheter dependent on being surrounded by liquid?  The IRRAflow ICP monitor is based on hydrostatic pressures requiring liquid around the side-holes of the catheter tip.  **IRRAflow device failure**  #1 Why did the IRRAflow show incorrect ICP values?  The side-holes of the catheter must be surrounded by liquid to evaluate the ICP  #2 Why do the side-holes have to be surrounded by liquid?  The IRRAflow ICP monitor is based on hydrostatic pressures requiring liquid around the side-holes of the catheter tip.  #3 Why was there no liquid around the catheter tip?  The catheter tip was surrounded by hematoma  **Diameter of catheter lumen**  #1 Why was the risks of occlusion related to the diameter of the IRRAflow catheter?  The inner diameter of the IRRAflow drainage line is 1.5 mm whereas the inner diameter of a standard 10 fr Silverline catheter is 1.9 mm.  #2 Why is the IRRAflow drainage line inner diameter smaller than the inner diameter of an EVD?  The IRRAflow catheter is a dual lumen catheter with an irrigation line and drainage line.  #3 Why is the risks of occlusion potentially higher in the IRRAflow catheter compared to a standard EVD?  Small lumen catheter is at significantly higher risks of occlusions than large diameter catheters  #4 Why does the irrigation of the IRRAflow not avoid occlusions?  The catheter is divided in two lumens and only the tip is a combined lumen which is held open by irrigation. The inner part of the catheter is separated, and the drainage line is therefore not held open by the irrigation.  **Aspiration method of the IRRAflow**  #1 Why could the occlusion be due to the aspiration method of the IRRAflow?  The IRRAflow performs suction instead of passive drainage as the drainage bag is placed from 0 to -103 cm below the patient causing a negative pressure gradient with accompanied suction.  #2 Why is suction needed when using the IRRAflow device  It is thought that when irrigation a volume of saline into the ventricles it should be followed by a suction mechanism to make sure of sufficient drainage.  #3 Why could suction cause occlusion  Suction can cause collapse of the ependyma around the catheter tip or by sucking the hematoma through the holes in the catheter tip. | **Most likely cause of event:**  Occlusion due to location of catheter tip in hematoma.  **Recommendation:**  Catheter should be functioning inside the hematoma and maybe be accompanied by tPA administration. The inner diameter of the drainage line in the IRRAflow catheter would favorably be increased. |
| Problem 3)  Event: SAE grade 3, displacement – tPA and irrigation into parenchyma  A control head CT revealed that the catheter had displaced into the parenchyma. With the tip of the catheter in parenchyma irrigation and tPA had been administrated directly in the parenchyma.  **Problem statement**  tPA and irrigation in parenchyma | The patient had a per protocol CT performed at day 4. The CT revealed that the catheter had displaced into the parenchyma. Due to the displacement irrigation in the parenchyma had occurred and further tPA had been administrated into the parenchyma.  Head CT showing tip in parenchyma (right) 25.05.22  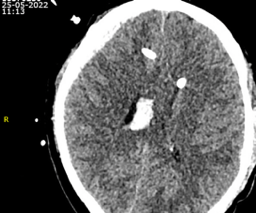 | Catheter displacement due to fixation method | **Catheter displacement due to fixation method**  #1 Why was tPA administrated directly into the parenchyma?  Due to catheter tip displacement into the parenchyma  #2 Why could the fixation method cause the displacement?  The IRRAflow is placed by tunneling and fixed using a fixation cap and sutures.  #3 Why are tunneled, suture fixated catheter more likely of displacing?  Catheter fixed by sutures are more likely to displace compared to bolted EVD as they are not as securely fastened as bolted catheters.  #4 Why is bolted EVDs more securely fastened?  The bolt is screwed through the cranium and the bolt is screwed until the catheter is securely fastened.  #5 Why is the IRRAflow not placed on a bolt?  The IRRAflow catheter lacs a metal enforcement enabling secure fastening of the bolt around the catheter to prevent constriction of the catheter. | **Most likely cause:**  Catheter fixation method with fixation cap and sutures  **Recommendations:**  The IRRAflow catheter should be compatible to bolted placement. |
| Problem 4)  Event: SAE grade 3, catheter replacement  The patient was still treated with the contralateral EVD after IRRAflow was removed.  ICP was found to increase, and the drainage was observed to be decreased. Due to semi occlusion of the catheter the catheter was replaced.  **Problem statement**  Catheter replacement due to occlusion | The ICP was observed to increase due to decreased CSF drainage. Brain debris and blot was seen in tubes of the EVD. Due to that the catheter was replaced using the same bolt and parenchymal canal. | Catheter occlusion | **Catheter occlusion**  #1 Why was the catheter replaced?  Due to malfunctioning with compromised CSF drainage.  #2 Why was the catheter malfunctioning?  Due to occlusion of the catheter  #3 Why was the catheter occluded?  Due to blood clots and brain debris  #4 Why did inflamed brain debris cause occlusion?  Inflamed brain debris are found to cause occlusion of catheters and seemed to be the reason for the occlusion. | **Most likely cause:**  Occlusion of the catheter  **Recommendations:**  Catheters should be left as short time as possible. |

| **Patient ID 16**  Enrolment September 2022   - ICH with break through - Graeb score 8 - Unilateral catheter treatment | | | | |
| --- | --- | --- | --- | --- |
| Problem 1)  Event: SAE grade 4, rebleeding  Prior to irrigation start after catheter placement the patient received a per protocol control head CT to ensure catheter placement in the ventricular system. CT revealed a worsening of IVH.  **Problem statement**  Rebleeding, worsening of IVH | The patient was admitted during the night and was randomized to IRRAflow treatment. The catheter was placed and further the patient had a hematoma in the posterior fossa removed surgically. Postop. CT scan showed worsening of the IVH. | Catheter placement and irrigation  Surgery in the posterior fossa and spontaneous rebleeding | **Catheter placement**  #1 Why could the catheter placement cause the worsening of the IVH?  Catheter placement is a surgical procedure which are always followed by a risk of bleeding.  #2 Why did it seem unlikely that catheter placement was the reason for the rebleeding?  The CT showed no sign of new hematoma in relation to the newly placed catheter  #3 why could irrigation potentially cause the worsening of the IVH?  Irrigation into parenchyma or in relation to small vessels could potentially cause the vessels to break and cause the worsening of the hemorrhage.  #4 Why did it seem unlikely that irrigation was the reason for the rebleeding.  The CT was made prior to irrigation start to ensure placement within the ventricular system.  **Surgery in the posterior fossa and spontaneous rebleeding**  #1 Why could the surgery in the posterior fossa cause worsening of the IVH?  Bleeding complications during surgery are very likely and could have worsened the IVH. However, the CT shows a well removed hematoma and no signs of bleeding during surgery.  #2 Why could the rebleeding be spontaneous?  A spontaneous rebleeding is very likely and could potentially have happened between the primary scan and the hematoma removal.  #3 Why is the spontaneous rebleeding more likely than a complication to the surgery in the posterior fossa?  The increase in amount of blood was found within the ventricular system. Mainly in the lateral ventricle and third ventricle. The postop. CT scan showed a well evacuated hematoma in the posterior fossa with only a small amount of blood in the fourth ventricle. | **Most likely cause of event:**  Spontaneous rebleeding  **Recommendation:**  None |
| Problem 2)  Event: SAE grade 3, displacement – irrigation into parenchyma  A control head CT was performed due to deficient awakening of the patient. CT revealed displacement of the catheter into the parenchyma which had caused irrigation into the parenchyma.  **Problem statement**  Catheter displacement and irrigation in parenchyma | Control head CT performed due to deficient clinical progress revealed displacement of tip into left caudate nucleus. Due to that irrigation had occurred into the parenchyma.  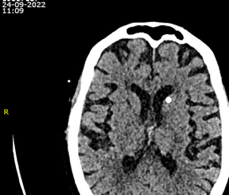 | Catheter displacement due to fixation method | **Catheter displacement due to fixation method**  #1 Why had irrigation in the parenchyma occur?  Due to catheter tip displacement into the parenchyma (caudate nucleus)  #2 Why was the catheter tip displaced?  Due to fixation method of the IRRAflow.  #3 Why could the fixation method cause the event?  The IRRAflow is placed by tunneling and fixed using a fixation cap and sutures. Catheter fixed by sutures are more likely to displace compared to bolted EVD.  #4 Why is bolted EVDs more securely fastened?  The bolt is screwed through the cranium and the bolt is screwed until the catheter is securely fastened.  #5 Why is the IRRAflow not placed on a bolt?  The IRRAflow catheter lacs a metal enforcement enabling secure fastening of the bolt around the catheter without occluding the catheter lumens. | **Most likely cause:**  Catheter fixation method with fixation cap and sutures  **Recommendations:**  The IRRAflow catheter should be compatible to bolted placement. |

| **Patient ID 17**  Enrolment October 2022   - SAH with break through - Graeb score 11 - Bilateral catheter treatment - Left lateral ventricle: EVD | | | | |
| --- | --- | --- | --- | --- |
| Problem 1)  Event: SAE grade 5, died  **Problem statement**  Patient passed away | Severe SAH and poor prognosis from the beginning. | Poor prognosis after severe hemorrhage | **Poor prognosis after severe hemorrhage**  #1 Why did the patient die?  Due to end of active treatment  #2 Why was the active treatment ended?  Due to very poor prognosis  #3 Why was the prognosis poor?  The patient had experienced a very severe SAH and was observed for 3 days without sedation with no sign of awakening.  #4 Why was there no sign of awakening? Vegetative state due to severe brain damage. | **Recommendations:**  None |

**Patient ID 18**

Enrolment January 2022	No SAEs observed

- ICH with break through
- Graeb score 6
- Unilateral catheter treatment

| **Patient ID 22**  Enrolment November 2022   - SAH with break through - Graeb score 4 - Bilateral catheter treatment - Right lateral ventricle: IRRAflow catheter - Left lateral ventricle: EVD | | | | |
| --- | --- | --- | --- | --- |
| Problem 1)  Event: SAE grade 4, contralateral EVD placement  The day after IRRAflow placement IRRAflow ICP values were very fluctuating, and no drainage was observed. ICP at control device increased to 30 mmHg.  **Problem statement**  ICP increase, no drainage and acute severe hydrocephalus | Control CT showed severe hydrocephalus and worsening of the IVH. The catheter tip was now placed in hematoma due to the rebleeding. An additional EVD was placed hyper acutely on the contralateral side for ICP control.  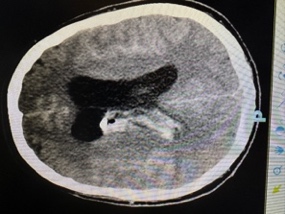 | Occlusion due to blood clots  Device failure  Diameter of catheter lumen | **Occlusion due to blood clots**  #1 Why was a contra lateral EVD placed?  Due to IRRAflow malfunctioning with no CSF drainage.  #2 Why was the IRRAflow malfunctioning?  The catheter tip was placed in hematoma.  #3 Why did the catheter localization in hematoma cause drainage stop  The catheter was occluded.  #4 Why did the catheter localization cause incorrect ICP values?  The side-holes of the IRRAflow catheter were placed in hematoma and not surrounded by CSF.  #5 Why was the tip of the IRRAflow catheter dependent on being surrounded by liquid?  The IRRAflow ICP monitor is based on hydrostatic pressures requiring liquid around the side-holes of the catheter tip.  **IRRAflow device failure**  #1 Why did the IRRAflow show incorrect ICP values?  The side-holes of the catheter must be surrounded by liquid to evaluate the ICP  #2 Why do the side-holes have to be surrounded by liquid?  The IRRAflow ICP monitor is based on hydrostatic pressures requiring liquid around the side-holes of the catheter tip.  #3 Why was there no liquid around the catheter tip?  The catheter tip was surrounded by hematoma  **Diameter of catheter lumen**  #1 Why was the risks of occlusion related to the diameter of the IRRAflow catheter?  The inner diameter of the IRRAflow drainage line is 1.5 mm whereas the inner diameter of a standard 10 fr Silverline catheter is 1.9 mm.  #2 Why is the IRRAflow drainage line inner diameter smaller than the inner diameter of an EVD?  The IRRAflow catheter is a dual lumen catheter with an irrigation line and drainage line.  #3 Why is the risks of occlusion potentially higher in the IRRAflow catheter compared to a standard EVD?  Small lumen catheter is at significantly higher risks of occlusions than large diameter catheters  #4 Why does the irrigation of the IRRAflow not avoid occlusions?  The catheter is divided in two lumens and only the tip is a combined lumen which is held open by irrigation. The inner part of the catheter is separated, and the drainage line is therefore not held open by the irrigation. | **Most likely cause of event:**  Occlusion due to location of catheter tip in hematoma.  **Recommendation:**  Catheter should be placed outside of the hematoma or if placed inside the hematoma be accompanied by tPA administration. |
| Problem 2)  Event: SAE grade 4, rebleeding  Control head CT shows severe worsening of hematoma in right lateral ventricle  **Problem statement**  Severe worsening of hemorrhage | Control head CT reveals rebleeding in the ventricular system. Graeb score increased from 4 to 6 when comparing to head CT from just after catheter placement. | Catheter placement and irrigation  Second aneurismal hemorrhage and coil procedure related complication | **Catheter placement and irrigation**  #1 Why could the catheter placement cause the worsening of the IVH?  Catheter placement is a surgical procedure which are always followed by a risk of bleeding.  #2 Why did it seem unlikely that catheter placement was the reason for the rebleeding?  The head CT performed just after catheter placement showed no sign of new hematoma in relation to the newly placed catheter  #2 Why could irrigation potentially cause the worsening of the IVH?  Irrigation into parenchyma or in relation to small vessels could potentially cause the vessels to break and cause the worsening of the hemorrhage.  #4 Why could the small vessels break?  The most optimal rate of irrigating using the IRRAflow has not been clarified. High rates of irrigation could potentially cause the small vessels to break.  #5 Why was irrigation the most likely reason for the rebleeding?  The rebleeding was found to be isolated to the right lateral ventricle and the third where the catheter was placed.  **Aneurysmal rebleeding**  #1 Why could the rebleeding be caused by a rebleeding from the aneurism?  The patient has suffered a SAH from a ACOM aneurism. Untreated aneurism is at high risk of re rupturing.  #2 Why was rebleeding from the aneurism less likely to have caused the rebleeding?  The location of hemorrhage was fare from the aneurism and further the aneurism was priorly to the rebleeding treated with endovascular coils.  #3 Why could the rebleeding be caused by a complication during coil procedure.  Risks of vessel perforation are present in endovascular treated aneurism patients  #4 Why was rebleeding due to a complication during the coil procedure less likely?  The location of hemorrhage was fare from the aneurism and no contrast was observed outside of the vessels. | **Most likely cause of event:**  Irrigation causing the hemorrhage  **Recommendation:**  Thorough investigation of safe and most optimal irrigation rates of the IRRAflow should be performed. |
| Problem 3)  Event: SAE grade 4, rebleeding  Control head CT showed severe worsening of hematoma in right lateral ventricle  **Problem statement**  Severe worsening of hemorrhage | Control head CT revealed a rebleeding in the ventricular system. The Graeb score increased from 6 to 8. | Catheter placement and irrigation  Second aneurismal hemorrhage and coil procedure related complication | **Catheter placement and irrigation**  #1 Why could the catheter placement cause the worsening of the IVH?  Catheter placement is a surgical procedure which are always followed by a risk of bleeding.  #2 Why did it seem unlikely that catheter placement was the reason for the rebleeding?  The head CT performed just after catheter placement showed no sign of new hematoma in relation to the newly placed catheter  #2 Why could irrigation potentially cause the worsening of the IVH?  Irrigation into parenchyma or in relation to small vessels could potentially cause the vessels to break and cause the worsening of the hemorrhage.  #4 Why could the small vessels break?  The most optimal rate of irrigating using the IRRAflow has not been clarified. High rates of irrigation could potentially cause the small vessels to break.  #5 Why was irrigation the most likely reason for the rebleeding?  The rebleeding was found to be isolated to the right lateral ventricle and the third where the catheter was placed.  **Aneurysmal rebleeding**  #1 Why could the rebleeding be caused by a rebleeding from the aneurism?  The patient has suffered a SAH from a ACOM aneurism. Untreated aneurism is at high risk of re rupturing.  #2 Why was rebleeding from the aneurism less likely to have caused the rebleeding?  The location of hemorrhage was fare from the aneurism and further the aneurism was priorly to the rebleeding treated with endovascular coils.  #3 Why could the rebleeding be caused by a complication during coil procedure.  Risks of vessel perforation are present in endovascular treated aneurism patients  #4 Why was rebleeding due to a complication during the coil procedure less likely?  The location of hemorrhage was fare from the aneurism and no contrast was observed outside of the vessels. | **Most likely cause of event:**  Irrigation causing the hemorrhage  **Recommendation:**  Thorough investigation of safe and most optimal irrigation rates of the IRRAflow should be performed. |

| Problem definition  Description of the event and formation of a problem statement. | Data gathering  Incident investigation: Collection of information regarding the event. | Causal factors  Causal factors causing the event | Root causes  The five-way analysis of root causes | Recommendations  Preventive action to prevent event from happening again |
| --- | --- | --- | --- | --- |
| **CONTROL GROUP** |  |  |  |  |

| **Patient ID 3**  Enrolment January 2022   - SAH with break through - Graeb score 12 - Unilateral catheter treatment | | | | |
| --- | --- | --- | --- | --- |
| Problem 1)  Event: SAE grade 5, died  **Problem statement**  Patient passed away | Very poor prognosis from a severe SAH. Active treatment was decided to be stopped after 3 days without sedation and found with only flexion extension. The patient dies expectedly. | Poor prognosis after severe hemorrhage. | **Poor prognosis after severe hemorrhage**  #1 Why did the patient die?  Due to end of active treatment  #2 Why was the active treatment ended?  Due to very poor prognosis  #3 Why was the prognosis poor?  The patient had experienced a very severe SAH and was observed for 3 days without sedation  The patient was extubated and died expectedly. | **Recommendations:**  None |

| **Patient ID 4**  Enrolment January 2022   - SAH with break through - Graeb score 11 - Unilateral catheter treatment | | | | |
| --- | --- | --- | --- | --- |
| Problem 1)  Event: SAE grade 5, died  **Problem statement**  Patient passed away | Very poor prognosis from a severe SAH. Active treatment is decided to be stopped the same day as admission due to very poor prognosis and the patient dies expectedly. | Poor prognosis after severe hemorrhage. | **Poor prognosis after severe hemorrhage**  #1 Why did the patient die?  Due to end of active treatment  #2 Why was the active treatment ended?  Due to very poor prognosis  #3 Why was the prognosis poor?  The patient had experienced a very severe SAH and was will sufficient treatment found to have an ICP of 50 with no response to any treatment actions. The patient was extubated and died expectedly. | **Recommendations:**  None |

**Patient ID 5**

Enrolment February 2022	No SAEs observed

- ICH with break through
- Graeb score 6
- Bilateral catheter treatment

**Patient ID 7**

Enrolment March 2022	No SAEs observed

- ICH with break through
- Graeb score 8
- Unilateral catheter treatment

**Patient ID 10**

Enrolment May 2022	No SAEs observed

- ICH with break through
- Graeb score 9
- Bilateral catheter treatment

**Patient ID 12**

Enrolment June 2022	No SAEs observed

- ICH with break through
- Graeb score 10
- Unilateral catheter treatment

| **Patient ID 13**  Enrolment June 2022   - ICH with break through - Graeb score 9 - Bilateral catheter treatment | | | | |
| --- | --- | --- | --- | --- |
| Problem 1)  Event: SAE grade 5, died  **Problem statement**  Patient passed away | Very poor prognosis from a severe cerebellar ICH with extension to the ventricles. MRI revealed infarction in the brain stem. Active treatment was decided to be stopped and the patient died expectedly. | Poor prognosis after severe hemorrhage. | **Poor prognosis after severe hemorrhage**  #1 Why did the patient die?  Due to end of active treatment  #2 Why was the active treatment ended?  Due to very poor prognosis  #3 Why was the prognosis poor?  The patient had experienced a very severe cerebellar hemorrhage with intraventricular extension and infarction in the brainstem. The patient was extubated and died expectedly. | **Recommendations:**  None |

| **Patient ID 15**  Enrolment August 2022   - ICH with break through - Graeb score 10 - Bilateral catheter treatment (bilateral EVD) | | | | |
| --- | --- | --- | --- | --- |
| Problem 1)  Event: SAE grade 3, Contralateral EVD placed  Through several days ICP lability followed by malfunctioning EVD with no drainage.  **Problem statement**  No drainage of CSF, EVD placed contralaterally | The patient had been experiencing fluctuating ICP and the drainage was observed to be fully compromised with no oscillation of the CSF in the EVD tube system due to occlusion. | Occlusion due to blood clots | **Occlusion due to blood clots**  #1 Why was a contra lateral EVD placed?  Due to EVD malfunctioning with no CSF drainage.  #2 Why was the EVD malfunctioning?  The catheter tip was placed in hematoma.  #3 Why did the catheter localization in hematoma cause drainage stop  The catheter was occluded.  #4 Why did the catheter localization cause occlusion?  The side-holes of the EVD catheter were placed in hematoma and not surrounded by CSF.  #5 Why was the tip of the EVD catheter dependent on being surrounded by liquid?  Blot clots are after coagulation very compacted and not able to be drained through the small lumen of an EVD. | **Most likely cause of event:**  Occlusion due to location of catheter tip in hematoma.  **Recommendation:**  Catheter should be placed outside of the hematoma or if placed inside the hematoma be accompanied by tPA administration. |

| **Patient ID 19**  Enrolment October 2022   - SAH with break through - Graeb score 10 - Bilateral catheter treatment (bilateral EVD) | | | | |
| --- | --- | --- | --- | --- |
| Problem 1)  Event: SAE grade 4, rebleeding  Rebleeding during coil treatment. During endovascular treatment of an aneurism, a perforation of the vessel happens with a more severe SAH following the complication.  **Problem statement**  Rebleeding | During the endovascular treatment of the patient’s aneurism a perforation of a vessel accidentally occurs with a more severe SAH following the complication. | Rebleeding from aneurism and endovascular treatment | **Rebleeding from the aneurism and endovascular treatment**  #1 Why could the rebleeding be caused by a rebleeding from the aneurism?  The patient has suffered a SAH. An untreated aneurism is at high risk of re rupturing.  #2 Why could the rebleeding be caused by a complication during coil procedure.  Risks of vessel perforation are present in endovascular treated aneurism patients  #3 Why was rebleeding due to a complication during the coil procedure most likely?  The location of hemorrhage related to the perforation and contrast was observed outside of the vessels. | **Most likely cause of event:**  Rebleeding due to perforated vessel during coil procedure  **Recommendation:**  None |

| **Patient ID 20**  Enrolment October 2022   - SAH with break through - Graeb score 8 - Unilateral catheter treatment | | | | |
| --- | --- | --- | --- | --- |
| Problem 1)  Event: SAE grade 3, ventriculitis  Elevated temperature from the 10^th^ of November. Primary suspicion of focus is CNS due to fluctuating GCS scores.  **Problem statement**  Fever with CNS as primary suspected infectious focus | Due to elevated temperature and fluctuating GCS a CSF sample was evaluated and found positive for coagulase negative staphylococci. Patient was treated with cefuroxime 3000mg x 3 | Catheter treatment  Hematogenous spread | **Infection related to catheter treatment:**  #1 Why was the infection related to catheter treatment?  Foreign objects are at risks of biofilm coating and are very bacteriophages.  #2 Why was the EVD at risk of causing infection? The EVD is silver impregnated which is shown to be less antibacterial compared to antibiotic coating.  **Hematogenous spread:**  #1 Why could the infection origin from hematogenous spread?  The patient had a positive urine sample  #2 Why was hematogenous spread less likely to have caused the positive CSF sample?  The positive sample of CSF was positive for a different bacterium than enterococcus faecalis.  #3 Why was an alternative primary infection not a possibility?  An alternative primary focus was not found or suspected | **Most likely cause of event:**  Catheter related infection  **Recommendation:**  Catheters should be left for as short time as clinically possible |
