## Supplementary material for "Safety, harm, and efficacy of IRRAflow^®^ versus external ventricular drainage for intraventricular hemorrhage: A randomized clinical trial": All supplemenatry: Supplementary 1ACTIVE Protocol v1 27012021.docx

**CLINICAL STUDY PROTOCOL**

**ACTIVE STUDY**

Use of **ACTIVE** Fluid Exchange to Treat Intraventricular Hemorrhage

**Protocol Number 47723569**

**27-01-2021 Version 1.0**

**Sponsored By Aarhus University**

**Supported with educational grant from IRRAS**

**Dansk titel: Brug af AKTIV væske udskiftning som behandling af intraventrikulær blødning**

**Contact Information Sheet**

| **Title:** | ACTIVE Study |
| --- | --- |
| **Protocol Number:** | TBD |
| **Device Class/ Study Phase:** | Use of Class II and III devices / Single Center Study |
| **Study Devices:** | IRRA*flow*® Active Fluid Exchange System |
| **Study Sponsor:** | Aarhus University with an unrestricted educational grant from IRRAS |
| **Study Sponsor Contact:** | Dr. Anders Rosendal Korshøj Aarhus University, Denmark  |
| **International Primary Investigator(s):** | Dr. Anders Rosendal Korshøj Aarhus University, Denmark |
| **Imaging Core Lab:** | TBD |
| **CRO and EDC Management** | Aarhus University and Aarhus University Hospital |
| **Protocol Version and Release Date:** | V.1.0 31.01.2021 |
| **Revision History:** | A |
| **Study Funding and Financial Support:** | Grant support from IRRAS, expected 3 million DKK over the study period |
| **Study IDs:** | Clinicaltrials.gov ID:  The Central Denmark Region Committee for Health Research Ethics Jr. Id: 77919 |

**Study Overview**

| **Study Device:** | IRRA*flow*® CNS System |
| --- | --- |
| **Title:** | Use of **ACTIVE** Fluid Exchange to Treat Intraventricular Hemorrhage |
| **Study Objectives:** | We hypothesize that active irrigation using the IRRA*flow* system will reduce the time needed for clearance of blood from the intraventricular space compared with passive drainage alone. Further, the study has been designed to confirm the IRRA*flow* system’s active irrigation will reduce the incidence of catheter occlusions, catheter-associated infections, treatment time, patient length of stay, and overall treatment cost when compared with passive drainage. Additionally, the information gathered from the IRRA*flow* and other procedural information can be utilized on other research projects, e.g. exploration of pressure/volume characteristics and related neuromonitoring. |
| **Study Design:** | Prospective, controlled, randomized, single-center, phase 2 study   - Study Type: Interventional - Allocation: Dual-Arm 1:2 randomization (intervention/control) - Endpoint Classification: Safety and Efficacy Study (phase 2) - Blinding: None - Masking: None - Purpose: Treatment |
| **Device Description:** | The IRRA*flow*® System is an active fluid exchange system in which intracranial pressure (ICP) monitoring, active controlled irrigation and passive drainage are combined in one system. The system consists of 3 items; control unit (i) and two sterile disposable parts: (ii) Intelligent Digital Cassette (Tube Set) and (iii) dual-lumen Catheter. |
| **Indications for Use:** | The use of IRRA*flow* CNS System is indicated when intracranial pressure monitoring is required and for externally draining intracranial fluid as a means of reducing intracranial pressure in patients where an external drainage and monitoring system is needed for ≤5 days. |
| **Regulatory Status:** | FDA 510(k) cleared - K171880  CE Mark – 12/19/2019 |
| **Treatment Population:** | Intraventricular Hemorrhage (IVH) |
| **Sample Size & Duration of Study:** | Expected 105 patients.  24-month study duration.  Interim analysis at 30 patients and 60 patients. |
| **Number of Sites:** | 1 |
| **Objectives and endpoints:** | Overall objective: To evaluate efficacy and safety of evacuation of intraventricular hematoma by Active External Ventricular Drainage (IRRA*flow*) compared to Passive External Ventricular Drainage (EVD).  The primary endpoint:   1. Time to clearance of ventricular blood as measured by head CT scan.   Secondary endpoints:   2. Rate of catheter occlusion. 3. Rate of catheter related infection. 4. Length of ICU stay 5. Rate of shunt dependency 6. Functional Status – Barthel Index, eGOS, and NIHSS at inclusion, 30 days, 3 months and 6 months. 7. Mortality rates at 30 days, 3 months and 6 months   Exploratory endpoints:   8. Duration of EVD in place 9. Procedure complications 10. Device related complications 11. Procedure success 12. Technical success and 13. Rate of revision procedures of the ventricular catheter 14. Rate of occurrence of repeat hemorrhagic events 15. Number of flushes required 16. Quality of Life utilizing the Stroke Impact Scale 17. Intensity of Critical Care Management – Hospital Days, ICU Days, ICP Management, Mechanical Ventilation, Pressors, Shunts, All infections and Pneumonia 18. Total cost of procedure |
| **Visit Schedule:** | Patients will be evaluated at baseline, during the procedure, and at discharge. |
| **Patient Selection Criteria:** | Patients who meet all the inclusion and exclusion criteria will be eligible for study participation. |
| **Inclusion Criteria:** | 1. Age >18 years of age 2. Intraventricular hemorrhage documented on head CT or MRI scan, no older than 72 hrs. 3. Need of cerebrospinal fluid drainage 4. Deterioration of consciousness or medical sedation at the time of enrollment 5. Indication for active treatment evaluated by the treating physicians 6. Signed informed consent obtained by patient or Legal Authorized Representative 7. Treatment possible within 72 hours of ictus 8. Use of validated anti-conception f fertile female participants in concordance with guidelines provided by the Danish Health and Medicines Authority |
| **Exclusion Criteria:** | 1. Patient has fixed and dilated pupils 2. Pregnant or nursing women (fertile female participants will be required to take a validated pregnancy test for evaluation of pregnancy) |
| **Target Enrollment Period/Study Duration:** | Enrollment period: 24 months  Follow-up period: 6 months  Total study period: 30 months |

**Statement of the Investigator**

- I agree to conduct the study in accordance with this protocol and to make no changes except when necessary to protect the safety, rights or welfare of patients. If such a change occurs, I will promptly inform the Sponsor of this event.
- I agree to personally conduct or supervise the study, and that all associates, colleagues and employees assisting me in the conduct of this study are informed of their obligations in meeting study commitments.
- I have read and understand the information in the Investigator’s Brochure and/or the Instructions for Use, including the potential risks, expected adverse events and potential side effects of the study itself and the product being studied.
- I agree to protect the rights of my patients and obtain informed consent from those who may participate in this study, in accordance with 21 CFR Part 56 and the requirements of my Institutional Review Board/Ethics Committee.
- I agree to report to the Sponsor adverse experiences that occur in the course of this study, in accordance with the study protocol requirements.
- I agree to maintain adequate and accurate records in accordance with study requirements and those records will be made available for inspection by the Sponsor, IRB/EC or regulatory bodies such as the U.S. Food & Drug Administration.
- I agree to control the distribution of study product provided to me in the course of this study and provide accurate accounting of the disposition of those products.
- I agree to provide my Institutional Review Board/Ethics Committee with all the information required to support both the initial and continuing review and approval of this study and I will not implement this study until such approval has been obtained. I agree to promptly report to the Institutional Review Board/Ethics Committee all changes in research activity and all unanticipated problems involving risk to patients or others.
- I agree to comply with all other requirements regarding the obligations of clinical investigators, as outlined in 21 CFR Part 812.100-150.

Investigator Name

Investigator Signature	Date

Institution Name

### 1. INTRODUCTION

Intracranial hemorrhagic conditions can rapidly cause brain damage and often considered life-threatening. Of these, Intracerebral hemorrhagic (ICH) is the most common type of hemorrhagic stroke and accounts for approximately 10-20% of all strokes *(1-3)* and associated with higher rates of morbidity and mortality than all stroke subtypes. Evidence suggests that the incidence of ICH also increases with advanced age *(4)*. The Global Burden of Disease 2010 study showed a 47% increase in the absolute number of hemorrhagic stroke cases (including ICH and subarachnoid hemorrhage) worldwide between 1990 and 2010 *(5).*

Intracerebral hemorrhagic stroke manifests through two main pathophysiologic mechanisms. The initial injury mechanism in ICH compresses the brain parenchyma by mass effect of hematoma, resulting in physical disruption of parenchymal architecture *(6).* Increased intracranial pressure due to expansion of the hematoma can affect blood flow, mechanical deformation, neurotransmitter release, mitochondrial dysfunction and membrane depolarization. As a result, neuronal injury in perihematomal area contains the edema and inflammatory environment by blood-derived factors *(7-9).* A secondary mechanism of brain injury is related to the clotting cascade, in particular thrombin, after endothelial damage and hemoglobin breakdown *(10-12).* Thrombin causes inflammatory cells to infiltrate the brain, proliferation of mesenchymal cells, formation of brain edema and scar tissue *(13).* Thrombin binds to protease-activated receptors *(1)* and activates the central nervous system microglia and complement cascade. As a result, multiple immune pathways are activated, which contributes to apoptosis and necrosis. Heme influx in neuron after endothelial damage leads to iron release and neuronal insult *(1, 14, 15).*

The most common clinical manifestation of ICH includes symptoms of headache, nausea and vomiting with increased intracranial pressure, or blood in the cerebrospinal fluid. Neurological deterioration is common before and during hospital admission and may indicate early hematoma enlargement or worsening of edema *(1).* Additionally, local intracranial effects (hydrocephalus and vasospams) and broader systemic consequences that affect pulmonary and cardiovascular systems *(16,17),* can pose considerable risk to patient health. Hematoma evacuation and decompressive craniectomy are options for treating elevated Intra Cranial Pressure (ICP) explained by the concept of preventing herniation, reducing ICP, and decreasing the pathophysiological impact of the hematoma on surrounding tissue.

Management of hemorrhagic patients is typically orchestrated by neurosurgeons and neuro-intensivists. Comprehensive care should include surveillance and monitoring of Intra Cranial Pressure, Cerebral Perfusion Pressure (CPP), and hemodynamic function. Furthermore, prevention of infection, complications of immobility through positioning and mobilization within physiological tolerance play an important role in optimizing outcomes after ICH.

There are multiple approaches to facilitating Cerebrospinal Fluid (CSF) drainage and monitor ICP. Routinely, intracranial pressure is measured by use of devices inserted into the brain parenchyma or cerebral ventricles. A Ventricular Catheter (VC) inserted into the lateral ventricle allows for drainage of CSF to help reduce ICP. Although CSF drainage is a vital sequence in patient management, there are reported risks including infection and limitations related to erroneous readings associated with current ICP monitors. Physicians lack the appropriate tools to employ active intermittent aspiration and drainage with continuous ICP monitoring.

The current clinical study is being initiated to evaluate the hypothesis that active irrigation by IRRA*flow*® will reduce the time needed for clearance of intraventricular blood from intraventricular space compared with passive drainage. Further, active controlled irrigation can improve catheter occlusion and infection rates compared with passive drainage.

### 2. STUDY DESIGN

#### 2.1 Description of the Study Design

The proposed study is a single center prospective, controlled, randomized trial to evaluate the efficacy and safety of evacuation of intraventricular hematoma by Active External Ventricular Drainage (IRRAflow) compared to Passive External Ventricular Drainage (EVD).

#### 4.2 Study Intervention and the Randomization Process

Randomization of the study will occur following enrollment upon the patient presentation to the emergency room. Upon initial diagnosis and enrollment, if it is determined that a CSF drainage is necessary, the patient will be randomized to either the IRRA*flow* with Active Fluid Exchange arm (intervention) or a standard practice EVD arm (control). The randomization will occur in a 1 to 2 fashion. This means, that every third case will be randomized to the intervention and two thirds to control. Drainage therapy with EVD or IRRA*flow* will commence for as long as it is deemed necessary by the treating physician. All patients enrolled in the trial will receive additional supportive and medical treatment by choice of the treating physician and in accordance with standard of care. Such treatment may include neurointensive care, neuromonitoring, and surgical or endovascular occlusion identified sources of intracranial hemorrhage, e.g. vascular anomalies, aneurisms, etc. Interventional treatment will be stopped in case of 1) patient exclusion from the trial, 2) ethical or medical safety contraindications for further interventional treatment determined by the Investigators.

Patient Presents to the Emergency Room with IVH

Screening and enrollment

Randomization 1:2

Intervention: IRRAflow Active Fluid Exchange Arm

Control: Traditional EVD Arm

**2.3 IRRA*flow* with Active Fluid Exchange Arm**

Patient identified and assessment conducted

Diagnostic CT of patient

Consent obtained

IRRAflow Catheter Inserted

IRRAflow TubeSet and Control Unit connected to the IRRAflow Catheter

CT Obtained to confirm placement of IRRAflow Catheter

Treatment Started

Treatment Algorithm and Metrics Recorded

CT performed at least every 48 hours to document effectiveness

Daily procedure and patient metrics recorded

CT and patient assessment utilized to determine completion of therapy

Final CT obtained with IRRAflow Catheter in place

Final procedure, patient and economic metrics recorded

Patient Discharge

Patient follow-up per standard of care

#### 2.4 Traditional EVD Arm

Patient identified and assessment conducted

Diagnostic CT of patient

Consent Obtained

EVD Inserted

CT Obtained to confirm placement of EVD

Treatment and Metrics Recorded

CT performed at lest every 48 hours to document effectiveness

Daily procedure and patient metrics recorded

CT and patient assessment utilized to determine completion of therapy

Final CT obtained with EVD Catheter in place

Final procedure, patient and economic metrics recorded

Patient Discharge

Patient follow-up per standard of care

#### 2.5 Duration of the Study

The enrollment period is estimated to be 24 months from the date of first patient enrolled. Patients will be followed for 6 months with expected trial termination after 30 months from initiation.

#### 2.6 Study Objectives

Overall objective: To evaluate efficacy and safety of evacuation of intraventricular hematoma by Active External Ventricular Drainage (IRRA*flow*) compared to Passive External Ventricular Drainage (EVD).


##### 2.6.1 Primary Endpoint

The primary endpoint is time to clearance of ventricular blood as measured by head CT scan.

##### 2.6.2 Secondary Endpoints

1. Rate of catheter occlusion.
2. Rate of catheter related infection.
3. Length of ICU stay
4. Rate of shunt dependency
5. Functional Status – Barthel Index, eGOS, and NIHSS at inclusion, 30 days, 3 months and 6 months.
6. Mortality rates at 30 days, 3 months and 6 months

##### 2.6.3 Exploratory Endpoints

1. Duration of EVD in place
2. Procedure complications
3. Device related complications
4. Procedure success
5. Technical success and
6. Rate of revision procedures of the ventricular catheter
7. Rate of occurrence of repeat hemorrhagic events
8. Number of flushes required
9. Quality of Life utilizing the Stroke Impact Scale
10. Intensity of Critical Care Management – Hospital Days, ICU Days, ICP Management, Mechanical Ventilation, Pressors, Shunts, All infections and Pneumonia
11. Total cost of procedure

##### 2.6.4 Study Outcome Definitions

- Stroke: An acute neurologic event with focal symptoms and signs, lasting for 24 hours or more.
- Major Stroke: A stroke defined as a new neurological event that persists for > 24 hours and results in a ≥ 4 point increase in the NIHSS score compared to baseline or compared to any subsequent lower score.
- Minor Stroke: A stroke that resolves completely within 7 days or increases the NIHSS by < 4 points.
- Neurologic death: Defined as a death which has been adjudicated by the treating physician to have directly resulted from a neurologic cause.

**2.6.5 Endpoint assessment and blinding**

Endpoints will be assessed by the Study Investigators and a statistical advisory service. Due to the nature of the intervention all assessments and treatment procedures will be unblinded.

### 3. TARGET POPULATION

The patient population for this study will be comprised of up to 105 patients with primary diagnosis of intraventricular hemorrhage or subarachnoid hemorrhage with intraventricular breakthrough

Patients who meet all the inclusion and exclusion criteria will be eligible for study participation.

#### 3.1 Study Criteria

Patient selection criteria are established according to clinical, radiologic, and neurologic components. Patients who meet all of the Inclusion Criteria and none of the Exclusion Criteria will be eligible for study participation.

A patient is considered to be enrolled after they have met all the inclusion and exclusion criteria for the study.

#### 3.2 Legal incompetence

Candidates for enrolment in the trial will typically be severely disabled by the acute neurological injury occurring due to hemorrhagic stroke. Most patients will be in a state of coma or medically sedated (and often intubated) leaving them incapacitated and unable to provide informed consent and the appropriate time of enrollment (see below)

#### 3.3 Inclusion Criteria

1. Age >18 years of age
2. Intraventricular hemorrhage documented on head CT or MRI scan, no older than 72 hrs.
3. Need of cerebrospinal fluid drainage
4. Deterioration of consciousness or medical sedation at the time of enrollment
5. Indication for active treatment evaluated by the treating physicians
6. Signed informed consent obtained by Experimental Representative (see below).
7. Treatment possible within 72 hours of ictus
8. Use of validated anti-conception f fertile female participants in concordance with guidelines provided by the Danish Health and Medicines Authority

#### 3.4 Exclusion Criteria

3. Patient has fixed and dilated pupils
4. Pregnant or nursing women (fertile female participants will be required to take a validated pregnancy test for evaluation of pregnancy)

#### 3.5 Sample size and statistical considerations

The analysis of the study endpoint will be performed on the ITT patient population. Subjects who are free from the events included in the primary endpoint composite, but who do not have complete follow-up, will be censored at the last time they were known to be event-free. Secondary outcome measures (subject-based unless otherwise specified) will be descriptive only; no formal statistical hypotheses will be tested. Secondary outcomes and demographics will be compared using t- test and chi squared tests where appropriate.

Proportions and confidence intervals will be produced for technical success, procedure success, and recurrence of hemorrhage. The confidence intervals will include a Bonferroni adjustment in order to maintain the family-wise coverage probability at 95%. Kaplan-Meier estimates will be used to calculate outcomes at 30 days and recurrence and neurologic death through 6 months.

### 4. STUDY PROCEDURES

For the purposes of the study the following are required:

- National Institutes of Health Stroke Scale (NIHSS) may only be performed by an individual certified to perform the NIHSS even if others may be qualified per state or local regulation. Any potential endpoint neurological symptoms will be assessed by the NIHSS. All NIHSS certifications must be kept up to date.
- Neurological exams will be performed by a NIHSS/mRS certified surrogate unless an actual neurologic event occurs, in which case the patient has to be evaluated by a neurologist.
- Other tasks may be performed by those individuals who are qualified by training and who have been designated by the site’s Principal Investigator to perform those tasks.

**4.1 Summary Data Collection and Schedule of Events**

Data will be collected on electronic case report forms at pre-procedure and during the procedure. A schedule of the study activities is provided below.

| **Visit (Window)** | Screening At Enrollment | Pre- Procedure  Within last 24hrs | Procedure | Prior to Hospital Discharge | After Discharge  3 and 6 months |
| --- | --- | --- | --- | --- | --- |
| Informed consent | X |  |  |  |  |
| Demographic Evaluation | X |  |  |  |  |
| Vitals (see notes below) | X*R | XR | XR | XR |  |
| Procedure Data |  |  | XR |  |  |
| mRS |  | X* |  |  | X |
| NIH Stroke Scale |  | X*R |  | XR |  |
| Glasgow Scale |  | X* |  |  | X |
| Barthel Index |  |  |  | X | X |
| SIS |  |  |  | X | X |
| EQ-VAS |  |  |  | X |  |
| ICH Score |  | X* |  |  |  |
| CT or MRI |  | XR |  |  |  |
| Concomitant Medications | XR | XR |  | XR |  |
| Adverse Events |  |  |  | XR |  |

| **NOTES:** |  |
| --- | --- |
| * | May be obtained at either screening or pre-procedure visit |
| ‡ | To be done only when clinically indicated |
| R | Standard of Care |
| Vitals | Vital signs should include Blood Pressure, Pulse and respiratory rate along with collection of height and weight. Temperature should be collected pre- and post- procedure at a minimum. |
| NIH Stroke Scale | Neurological assessments should be done by a physician or research personnel certified in the administration of NIHSS |
| Standard of Care | For the purposes of this study protocol, whatever neuro exams and procedural related parameters that need to be documented and assessed. |

To Read: History, Physical Examination, and Workup of the Patient With ICH – Refer to AHA-ASA Guideline for the Management of Spontaneous Intracerebral Hemorrhage_Hemphill_Stroke 2015

#### 4.3 First Contact and the Informed Consent Process

##### 4.3.1 Enrolment without prior consent

Given the acute nature of intraventricular hemorrhage and the associated neurological deterioration (e.g. due to coma, loss of consciousness, other severe neurodeterioration, or sedation/intubation), the majority of candidates for enrollment in this trial, will be considered incompetent and unable to provide written consent for trial participation. In addition, the experimental intervention and the standard intervention (control), should be initiated as soon as possible upon diagnosis of intraventricular hemorrhage in order to evacuate blood and reduce the intracranial pressure as these conditions can be immediately critical and life-threatening. In these cases, drain placement (IRRA*flow* or standard drain treatment) is standard practice and serves to evacuate blood and cerebrospinal fluid and thereby induce a life-saving decompression of the intracranial compartment. Delayed drain placement can cause prolonged intracranial hypertension leading to global brain hypoperfusion and severe secondary brain damage. Therefore, it is imperative, that potential participants are enrolled at first possible notice upon diagnosis to ensure swift treatment in due time. If immediate inclusion is not done, it will not be possible to enroll the patients at a later point in time, as they will have already received standard drainage therapy. For these reasons, it will generally be necessary to include patients in the trial without their prior informed consent in accordance with applicable law and ethical guidelines on “acute clinical trials”. In these instances, the trial investigators will obtain proper written consent from the patient or a Legal Authorized Representative upon first possible notice in accordance with §§3-5 in the Danish Law on Health Research Ethics (Komitéloven). In addition, Informed Consent will be given for each enrolled participant by a Legal Authorized Representative (LAR) in accordance with §12 of the Danish Law on Health Research Ethics. The LAR can be a near relative as defined in the applicable law or potentially a medical doctor who will represent the patients interests and who will be independent of the interests of the Principal Investigator and the trial *per se*. In cases where the LAR is a medical doctor, the LAR will be a person who is who is neither superior nor subordinate to the Principal Investigator. Based on his/her knowledge of the patients health and current condition as well as the written patient information material and oral information, the LAR will evaluate whether it is permissible and in the patients best interest to participate in the trial. Informed consent from the LAR may be given tentatively by means of oral information, e.g. by phone. In such cases, however, the written informed consent should always be obtained afterwards. The LAR may be any medical doctor with knowledge about the patients condition. As an example, the LAR will be a medical doctor on shift at the time of the candidate’s hospital admission and enrollment, e.g. an anesthesiologist, emergency doctor, neurosurgeon or neurologist. The trial protocol and written patient information will be made immediately available to the LAR.

Following the initial LAR consent (§12), subsequent informed consent will be obtained from the patient or the LAR in accordance with the Danish Law on Health Research Ethics §4.3 (see below).

##### 4.3.2 Subsequent Informed Consent Proces

Upon immediate enrollment in accordance with the above (§12) a subsequent informed consent will be obtained from the patient, the patient’s LAR. In these instances, patients or their LARs will be made aware of the experiment (first contact) at first possible notice. A written informed consent form shall be obtained from and signed by each potential patient (Patient) or LAR and by the Principal Investigator (or designee) according to the established procedures at the Investigation Site and any applicable law. Written Information regarding the study shall be handed to each Patient or LAR, per IRB/EC policy and corresponding oral information shall be given to each Patient or LAR in a language and level of complexity that each Patient/LAR will understand. Patients or LAR’s shall have sufficient time (at least 24 hours) to discuss and understand the rationale of participation in the study with the Principal Investigator (or designee) before potentially making a decision regarding participation. At the candidates will or the will of the LAR this decision can, however, be made immediately in relation to the information meeting. All questions or concerns shall be addressed to the complete satisfaction of the Patient or LAR prior to a final decision on enrollment in the study. No Patient shall not be coerced, persuaded or unduly influenced into participation into the study. If the Patient or LAR is unable to read the informed consent form, the Principal Investigator (or designee) shall read the consent form in full, prior to signing, and make witness to the consenting process. For non-English speaking patients, an interpreter will be provided if available by the site and both interpreter and the Patient or LAR will sign the ICF.

Information will be given by the Sponsor, the Investigators, the Co-investigators, or clinical staff members, who have been properly trained in the trial protocol. Meetings will be held in a quiet and peaceful atmosphere in a regular clinical consultation room at the local site’s enrolling department. The right for participants to be accompanied by an assessor to the oral information consultation will be stated in the written information and at initial contact. Fertile female candidates will be made aware of the requirement for the use of validated anti-conception throughout the entire trial period upon first contact.

##### 4.3.3 General Considerations

The informed consent forms (ICF) that is utilized for the study shall be reviewed and approved by the Institutional Review Board (IRB)/Ethics Committee (EC) at each Investigation Site and the Sponsor. The signed informed consent form shall be retained in the study files. Any amendments made to this protocol, patient information and the patient consent form must be approved by the IRBs/ECs.

#### 4.3 Pre-Screening/Screening

All patients who sign an ICF will ONLY be considered entered into the screening phase of the study but not enrolled yet. A patient screening log will be maintained at the study site. For patients who did not meet eligibility criteria after signing informed consent, the reasons for exclusion will be documented on the patient screening log. CT or MRI brain imaging is required for all patients to document the diagnosis.

Consent and enrollment will include permission to pass on health data to the project and its faculty members including clinical staff, academic staff, and organizations (including IRRAS) participating in the trial as well as relevant authorities and monitoring organizations. Health information will include information from the participant’s patient record and all recorded data. This information will be used to evaluate suitability for enrolment and also to evaluate the clinical status of participants prior to and during the experiment and assess and ensure the quality of trial execution. Consent for trial participation will be noted in the patient record. Original signed consent forms will be stored and protected. Patients will be made aware that consent can be withdrawn at any time.

#### 4.4 Procedure

- Patient presents into the emergency room with symptoms or findings that indicate need for drainage of CSF.
- The diagnosis of IVH is documented on head CT or MRI scan.
- Patient is identified as a potential candidate for the ACTIVE study.
- Upon assertion that the patient is a potential candidate for the ACTIVE Study, the patient will be randomized to either the IRRA*flow* with Active Fluid Exchange arm or the traditional EVD arm. The randomization will occur in a 1 to 2 fashion. This means, that every third case will be randomized to the IRRA*flow* with Active Fluid Exchange arm.
- Further assessment of the patient will be done to ensure they fit the inclusion criteria.
- If the patient fits the inclusions criteria, a consent will be obtained and the patient will be officially enrolled into the ACTIVE study.
- Treatment will be based upon the treatment path the patient is randomized to.
- Treatment will occur based on the protocol.
- Treatment will be completed per discretion of the physician.

#### 4.5 Post-Procedure

Patient will be required to stay in the hospital following the procedure. The following will be completed between 24-48 hours post procedure or prior to discharge, whichever comes first (unless otherwise indicated):

- Vital signs
- NIHSS
- Adverse events assessment
- Assess concomitant medications

#### 4.6 Follow-Up and Response Assessment

At this time, a scheduled follow-up patient visit is not required.

#### 4.7 Patient Withdrawal and Lost to Follow-Up

A patient or a patient’s LAR may elect to withdraw from this clinical study at any time. The patient or the patient’s LAR should notify the Investigator of the request to withdraw. The Investigator should encourage patients to return for all required follow-up visits and request that they return for the withdrawal visit. The Investigator may also withdraw the patient from the clinical study at any time based on his/her medical judgment.

All patients who withdraw from the study should complete an end of study visit. No further visits are required by the patient once the end of study visit is complete. A patient has the right to withdraw from the trial at any time and for any reason without prejudice to his or her future medical care by the physician or the institution. Trial withdrawal by a patient or their LAR specifically means withdrawal of consent from further participation in the trial. Patients who withdraw consent after enrollment will be evaluated to the time of withdrawal, and withdrawal of consent precludes any further trial-related treatment or data collection. LAR may also withdraw consent. At a minimum, every effort should be made to document patient outcome at the time of trial withdrawal.

All patients are expected to continue in the trial through the final follow-up assessment or until IRRAS notifies the Investigator in writing that further follow-up is no longer required, except in the event of death or upon the patient's (or LAR's) written request for early withdrawal from the clinical trial. A copy of this request should be forwarded directly to IRRAS for documentation.

The Investigator should encourage patients to return for all the required follow-up visits. These measures are important as the clinical study objectives may not be realized if a large number of patients are lost to follow-up (LTFU).

A patient will be considered LTFU and discontinued from the study once they have missed the 30-day follow-up visit. The Investigator must complete the appropriate CRF (Study Completion Form) documenting the patient’s withdrawal or discontinuation from the clinical study. The patient will be considered withdrawn once the Sponsor receives the respective CRFs.

The Investigator may also withdraw the patient from the clinical study at any time based on the Investigator’s medical judgment. The study Monitor will ensure that patient lost to follow-up and withdrawals are documented properly, discussed with the Investigator, and reported to the Sponsor.

#### 4.8 Patient Study Completion

A patient has completed the study when he/she has completed treatment. Any patient that does not complete these requirements due to voluntary withdrawal, physician withdrawal, lost to follow-up, death, or any other reason will be considered as an early withdrawal.

### 5. STUDY ADMINISTRATION AND MONITORING PROCEDURES

#### 5.1 Requirements Prior to Initiation of the Study

The use or disclosure of all protected health information will comply with the GDPR and adhere to strict professional standards of confidentiality.

#### 5.2 Device Training

All investigators and co-investigators will undergo standardized training in the use and operation of the IRRA*flow* device prior to study participation.

Physician device training will be provided by a qualified IRRAS Associate or designee. The minimum requirements for physician device training include - Didactic sessions to cover the directions for use, preparation of IRRA*flow* system and procedure planning.

The study personnel at each study site will undergo protocol training and a site initiation visit. Site training and initiation will be documented, and the records maintained by the site and study Sponsor.

#### 5.3 Sponsor Representatives

IRRAs representatives may be present during study procedures to provide technical assistance to the Investigator in the use of the device. The activities of these representatives will be supervised by the Investigator.

#### 5.4 Source data, data handling and protection of participant integrity

All data will be handled in accordance with *the Danish Act on Processing of Personal Data, the General, Data Protection Regulation (GDPR) of the European Union (EU)* and other relevant legislation. Data will be treated anonymously (encoded) in analyses and publications. Written consent for participation in the trial will include permission to handle, convey and pass on health-related information to the Sponsor, Sponsor representatives, and monitoring authorities from the participant’s patient record, including information and raw data concerning current and prior health, diagnoses, medical treatment, laboratory analyses, blood work, imaging data, electrophysiological data, treatment information, diagnostic procedures, as well as purely private matters and other confidential information, as part of safety evaluation, patient selection, quality control and monitoring. For these purposes, data may also be extracted from the electronic patient record. Consent for participation, clinical treatment and evaluation, adverse events and other issues of clinical relevance will be noted in the patient record. Personal health information, which is necessary for proper technical support by the IRRA*flow* technicians (e.g. name, age, contact information, diagnosis, CT scans), will be shared with IRRAS.

Data will be collected on source documents and input into the electronic data collection (EDC) system via electronic Case Report Forms (eCRFs). The Investigator will be responsible for the accuracy and completeness of the eCRFs. When submitted electronically, CRF completion will be tracked by the user name and password of the Investigator or designee. All electronic systems are validated according to GDPR and Good Clinical Practice standards.

#### 5.4 Monitoring and quality control

The project will comply with international guidelines good clinical practice (ICH-GCP and DS/EN ISO 14155:2020. Clinical investigation of medical devices for human subjects - good clinical practice) and be monitored by the local contract research organization (CRO). The Monitor shall assure that the investigators are complying with the signed investigator agreement, the Clinical Investigation Plan/Protocol, appropriate regulations and any conditions of approval imposed by the Investigation Site’s IRB/EC. The level of monitoring will be described in a separate monitoring plan. Source data, verification of consent, and handling of adverse events and near-miss incidents will be subject to sampled monitoring. Full access to all source data incl. patient records, will be granted in relation to trial inspection by the relevant national competent authorities. Experimental “waivers” from the protocol will be registered in the project trial master file and reported to the relevant authorities. Changes to the protocol will be submitted for approval by the relevant authorities in accordance with GCP guidelines and regulations.

#### 5.5 Records

- The Principal Investigator shall maintain accurate and complete records relating to the study. These records shall include Clinical Investigation Plan/Protocol, IRB/EC records, records of receipt, use and disposition of IRRA*flow* devices.
- The Principal Investigator may withdraw from the responsibility to maintain records for the time required by transferring custody to another person who will accept responsibility for them.
- The investigator is responsible for ensuring that data are properly recorded on each patient's source document, eCRFs, and other related documents. An investigator who has signed the protocol signature page should personally sign the eCRFs (as indicated in the eCRF) to ensure that the observations and findings are recorded on the eCRFs correctly and completely.
- Data will be maintained in a sponsor-approved central data repository.

### 6. MANAGEMENT OF ADVERSE EVENTS


#### 6.1 Adverse Event (AE)

Any unfavorable and unintended sign (including laboratory findings), symptom or disease that occurs to a patient resulting in a change in normal baseline health and while enrolled in this clinical study. Medical conditions that exist at study enrollment are not considered an AE unless the condition worsens after use of the study device.

#### 6.2 Serious Adverse Event (SAE)

Serious Adverse Event is an adverse event that:

- led to death
- led to serious deterioration in the health of the subject, that either resulted in –
  - a life-threatening illness or injury, or
  - a permanent impairment of a body structure or a body function, or
  - in-patient or prolonged hospitalization, or
  - medical or surgical intervention to prevent life threatening illness or injury or permanent impairment to a body structure or a body function structure

Note: Planned hospitalization for a pre-existing condition, or a procedure required by the study protocol, without serious deterioration in health is not considered a serious adverse event.

#### 6.3 Unanticipated Adverse Device Effect (UADE)

An unanticipated adverse device effect is any serious adverse effect on health or safety or any life-threatening problem or death caused by or associated with a device, if that effect, problem or death was not previously identified in nature, severity or degree of incidence in the investigational plan or IDE application (including a supplementary plan or application), or any other unanticipated serious problem associated with a device that related to the rights, safety or welfare of patients.

#### 6.4 Anticipated Adverse Events

Anticipated Adverse Events that are inherent to Intracranial fluid drainage procedure and expected to occur in most subjects for a projected duration, according to the medical opinion of the investigator, may be considered unavoidable. Such events include, but are not limited to, those listed in Table X below. These adverse events constitute expected clinical observations and should not be reported to the sponsor during this study but shall be reported to the Investigation Site’s IRB/EC per the IRB’s policy**.**

**Table X: Anticipated Adverse Events and Reporting Time Frame**

| **Description of the Event** | **Time Frame from the Index Procedure** |
| --- | --- |
| Blood electrolyte disturbances  (e.g. shift in sodium or potassium levels) | Within 48 hours |
| Anesthesia-related nausea and/or vomiting | Within 24 hours |
| Anesthesia-related diarrhea or constipation | Within 48 hours |
| Low-grade fever (< 100° F or < 37.8° C) | Within 48 hours |
| Back pain related to laying in bed | Within 48 hours |
| Incisional pain (pain at access site) | Within 72 hours |
| Sleep problems or insomnia | Within 72 hours |
| Mild to moderate bruising or ecchymosis | Within 168 hours |

If a complication occurs that is not on the list of known, potential complications and the investigator believes that the complication is a potential UADE, the site should immediately contact the Sponsor to determine reporting requirements

#### 6.5 Registration and reporting of adverse events and near-miss incidents

AE reporting procedures will comply with “guidelines on medical devices. Clinical investigations: serious adverse event reporting” MEDDEV 2.7/3”, and “Reporting of Adverse Reactions in Clinical Trials”. All SAEs and relevant AEs will be reported in scientific publications. Planned hospital admissions or interventions unrelated to the trial will not be registered as AEs. Possible causal relationship with the intervention will be considered carefully by the principal investigator for all AEs and near-miss incidents.

For this post-market study, only AE/SAEs that are related to the procedure, device or a change in neurological status should be captured on the appropriate electronic case report form (eCRF) for that visit and documented in the patient’s medical record. The Investigator at each Site is ultimately responsible for reporting AEs to the Sponsor.

Adverse event reporting begins once the patient is enrolled in the study. All adverse events should be reported from enrollment through study completion/discontinuation. Any events occurring between consent and enrollment should be documented in the patient’s medical history.

A diagnosis should be provided whenever possible. If unable to provide a diagnosis, report the symptoms as separate events. Adverse Events should be reported using the full name without abbreviations or narratives.

Adverse events with an outcome status of “Ongoing” should be assessed at each follow-up evaluation to determine if the event has resolved.

Adverse events ongoing at study completion/discontinuation should be left as “Ongoing” on the AE case report form.

The common terminology criteria for adverse events (CTCAE) will be utilized for Adverse Event (AE) reporting. The CTCAE v.4.0 incorporates certain elements of the MedDRA terminology.

| Table X: Adverse Event and Device Malfunction Reporting Times | | |
| --- | --- | --- |
| **CLASSIFICATION** | **REPORTING TIME** | **TYPE OF REPORT** |
| Unanticipated Adverse Device Effects (UADE) | Within 24 hours of learning of the event, no more than 3 working days from learning of event | Adverse Event Reporting Form to be submitted to the sponsor, CRO and IRB |
| Serious AE (SAE) | Within 24 hours of learning of the event, no more than 3 working days from learning of event | Serious Adverse Event Reporting Form to be submitted to the sponsor, CRO and IRB |
| Non-serious AE | Within 5 working days of the event | Adverse Event Reporting Form in EDC |
| Device Malfunctioning with or Without Adverse Event * | Within 5 working days of learning of the event | Investigational Clinical Device Product Complaint and Return Form to be submitted to the sponsor, CRO and IRB. In case of death, it should be submitted to the FDA, Sponsor, CRO and IRB. |

*A “device malfunction” is defined as a device that fails after introduction into the patient.


#### 6.6 Adverse Event Relationship

Each reported AE will be assessed by the Investigator for its primary suspected relationship to the IRRA*flow* use for its intended procedure

- **Device-Related**

If the functioning or characteristics of the studied devices caused or contributed significantly to the adverse event, the adverse event would be suspected as primarily related to studied device.

- **Study Procedure-Related**

If the procedure (and not the device) caused or significantly contributed to the adverse event, the adverse event would be suspected as primarily related to the procedure.

- **Medication-Related**

If the adverse event was a result of a medication taken as part patient treatment, the adverse event would be suspected as primarily related to the medication.

- **Disease-Related**

If the adverse event was a result of the underlying disease progression for which the study procedure is being performed, and not the device or procedure, the adverse event would be suspected as primarily related to the disease.

- **Not Related**

If an adverse event cannot be attributed to the device, procedure, medication, or disease, it will be reported as “Not related”.

- **Unknown Relationship**

If the relationship of the adverse event to the device, procedure, medication, or disease cannot be determined, it will be coded as “Unknown”.

#### 6.7 Patient Death

If a patient dies while participating in the study, the cause of death will be reported as a serious adverse event and “death” reported as the event outcome on the AE eCRF. If the patient has other ongoing adverse events at the time of death, the outcome status of those events should be reported as “resolved” on the AE case report form.

#### 6.8 Reporting of Adverse Events by Patients

All patients shall be instructed to report any adverse events to the Principal Investigator or Study Coordinator at hospital discharge and during the course of the study.

#### 6.9 Device Malfunction

A “device malfunction” is defined as a device that fails to function as intended. If IRRA*flow* system does not meet the specific performance requirements, per the provided Instructions for Use, the issue/incident must be reported. Another IRRA*flow* device shall be available to the physician to complete the procedure.

If the device malfunction results in an adverse event, fill out a Product Complaint Form and submit it to the sponsor.

### 7. ETHICS AND REGULATORY CONSIDERATIONS

#### 7.1 Subject Information and Consent Procedures

The Investigator, study coordinator or any other member of the research team approved by the IRB/EC will obtain written informed consent from the patient or his/her LAR before subjecting her/him to study assessment procedures. The Subject will be clearly informed that the principal investigator, study coordinator and study monitor of the coordinating site will have access to personally identifiable information for the purposes of monitoring data against source documentation. However, only de-identified data is entered into the study database.

Each site participating in this study will have their own consent form that must be approved by their IRB/EC. Only the local participating site consent form should be signed prior to enrollment and treatment. The original, signed and dated ICF should be retained in the Subject’s study records, and a copy provided to the Subject. Documentation of the informed consent process should also be retained in the subject’s study records.

#### 7.2 IRB/EC Approval

The Investigator or the study coordinator is responsible for submitting the study protocol and any changes issued by the Sponsor during the course of the study for IRB approval prior to any Subject enrollment and amendments taking effect as well as obtain renewals at periods determined by the IRB/EC for the duration of the study.

#### 7.3 Publication Plan

The Sponsor will register the study and post results on the clinicaltrials.gov registry as required by the International Committee of Medical Journal Editors (ICMJE) member journals and applicable U.S. laws and regulations. It is the intent of the sponsor that the results of this study will be submitted for a professional publication. The Sponsor will establish a publications committee that will review the results and develop publications during and at the completion of the study.

#### 7.4 Risks and safety precautions

General safety considerations concerning IRRA*flow* therapy are outlined in the related technical and safety material provided by IRRAS. In general, IRRA*flow* is a CE marked and US FDA approved technology, which is regarded a safe and recognized treatment for IVH. We expect the trial to produce benefit for society as a whole, in the sense that it will provide important information about a potential new and improved treatment opportunity with expected benefit for future IVH patients. In addition, we expect that the trial will provide individual benefit for enrolled patients, in the sense that 2/3 of the cohort will receive standard of care, while the remaining 1/3 will receive experimental treatment which can effectively function in a similar fashion to standard of care EVD but provide further active treatment option on a rational basis in the form of active clot removal.

##### 7.4.1 Contraindications

The IRRA*flow*® Active Fluid Exchange System is not suitable for lumbar drainage.

The use of the Control Unit is contradicted when trained personnel to supervise monitoring and drainage are not available.

The Control Unit is contraindicated for use in a Magnetic Resonance (MR) environment. Refer to the IRRA*flow* Catheter IFU for MR environment use.

##### 7.4.2 Warnings

- Only medical personnel with training and experience in neurosurgical medical care may perform treatments involving this Device. Use in any other way may potentially harm the patient and/or the user.
- Only IRRA*flow* Intelligent Digital Cassette and IRRA*flow* Catheter may be used together with IRRA*flow* Control Unit. Using other components can injure patients.
- To reduce the risk of interference from outside sources, avoid using the IRRA*flow* Control Unit and IRRA*flow* Intelligent Digital Cassette near strong sources of electromagnetic radiation (e.g. diathermy equipment, MRI).
- The patient may not touch the Control Unit during treatment. The treatment may be disturbed if the patient mistakenly touches any part of the equipment.
- No other components than USB memory sticks may be inserted into the USB memory slot in the IRRA*flow* Control Unit. Erroneous use could potentially endanger the integrity of the Control Unit.
- Treatment may not be conducted if the surrounding temperature or the atmospheric pressure exceeds any of the limits stated in the manual.
- ICP measurements are not reliable during defibrillation and necessary precautions need to be made in such an event.
- The equipment is not intended for use in oxygen rich environments or in the presence of flammable anaesthetic mixtures or other flammable gases.
- Modification or disassembly of the Control Unit is not permitted. Unauthorized modifications to the Control Unit can cause a malfunction resulting in serious patient injury, damage to internal circuitry or electric shock.
- Explosion Hazard: Do not use in the presence of flammable materials (e.g., anesthetics, solvents, cleaning agents and endogenous gases).
- Electrical Shock Hazard:
  - Use only IRRAS approved power supplies listed in Recommended Accessories and Reordering Information section.
  - Use of another power supply may not provide electrical isolation from supply mains and protection against electrical hazards.
- Do not remove side, front or rear panels. Contact Technical Support for service and repair.
- The IRRA*flow* Catheter is not suitable for lumbar introduction.
- The IRRA*flow* Intelligent Digital Cassette and IRRA*flow*® Catheter is not to be reused, reprocessed or re-sterilized when open but unused.

##### 7.4.3 Precautions

- The Catheter must not be connected to the IRRA*flow* Control Unit while setting up the Control Unit for treatment. This could potentially harm the patient.
- There is a risk of the user getting pinched when moving the Control Unit up or down. Use care when performing these actions.
- IRRA*flow* Intelligent Digital Cassette and IRRA*flow* Catheter are single-use components. Using the same component for multiple treatments can potentially harm the patient.
- The IRRA*flow* Catheter shall be unpacked and prepared in a sterile area and in a sterile manner.
- To avoid contamination, the IRRA*flow* Intelligent Digital Cassette and IRRA*flow* Catheter are to be handled with care when being attached. Special care should be taken with the Catheter, and the connection of the Intelligent Digital Cassette to the Catheter and the connection of the fluid drainage bag.
- Precautions must be taken when changing an empty drainage bag for a new bag to prevent patient infections.
- Precautions must be taken when disconnecting the IRRA*flow* Catheter from the IRRA*flow* Intelligent Digital Cassette to prevent patient infections.
- Only Irrigation fluids specified in this manual can be used when conducting treatments with the IRRA*flow* Active Fluid Exchange System. A completely new and sterile irrigation bag must be used for each new treatment.
- In order to have correct ICP measurements, and thus properly set pressure alarm levels, the 0 point of the control unit must always be aligned with the Catheter’s tip position intracranially, which corresponds to the patient’s external auditory meatus or top of the eyebrow. Care should be taken when moving the patient in the vertical axis in order to readjust the height of the control unit before restarting treatment.
- IV pole and patient bed wheels are to be locked during the treatment. Care should be taken when moving the patient.
- Set high and low ICP alarm limits before starting treatment according to the treating physician recommendation.
- Always follow the instructions for cleaning and disinfection found in this User Manual. If these instructions are not followed, the unit risks being damaged, and/or the patient and the user may be exposed to contaminated parts.
- If the IRRA*flow* Control Unit, IRRA*flow* Intelligent Digital Cassette or IRRA*flow* Catheter is used in a way that contradicts the intended use or by individuals who are not medical personnel with training and experience in neurological/ neurosurgical medical care, then this could result in injury to the patient and/or the user.
- Over-drainage of intracranial fluid may cause ventricular collapse and injury to the patient. The Catheter may be occluded by ventricular collapse. Always monitor drainage progress by checking the drained volume in the drainage bag.
- Never pour liquids on any part of the IRRAflow Control Unit. If this occurs, dry off with a clean cloth.
- Always follow the preventive maintenance instructions for the IRRAflow Control Unit.
- No tools need to be, nor should be used when handling the IRRA*flow* Control Unit. All attempts to open or modify the unit involve risks to the user and potentially to the patient.
- Only accessories delivered with the unit or provided by IRRAS or an IRRAS official distributor may be used. Using accessories from third parties may involve a safety risk and voids any warranty.
- Take USB precautions when using the USB contact.
- To avoid electric shock, this equipment must only be connected to a main with protective earth.

**7.5 Financial aspects**

The project is funded by the following institutions and amounts:

- IRRAS will support the trial with a grant. The amount is still to be defined. At present time we expect to receive approximately 3 million DKK from IRRAS over the project period

Furthermore, the project is launched on the initiative of the Sponsor but supported financially by IRRAAS. Funds will cover costs associated with the trial, including running costs, salaries, legal handling, and treatment expenses. The Sponsor/Principal Investigator is compensated with external funds unrelated to IRRAS. There will be no financial compensation for patients participating in the project.

**7.6 Insurance**

Patient insurance related to inflicted injuries due to malfunction of the IRRA*flow* device is provided by the manufacturers. For Danish patients, all other injuries are covered by the Danish Patient Compensation Association. For non-Danish participants treated at non-Danish sites, all other injuries, i.e. not related to malfunction of the IRRAf*low* device, will be covered by the local sites or their governing institutions. It will be required, that any participating site can document such insurance coverage

**7.7 Samples of biological material**

Blood samples will be taken as part of the standard clinical evaluation and treatment. All sampled material will be destroyed immediately after analysis.

### 8. END OF THE STUDY


#### 8.1 Expected Study End

The study will close once the desired number of patients has been enrolled and follow-up completed for all patients. This should take approximately 30 months.

#### 8.2 Early Study End

The study may be terminated early for any of the following reasons:

- New findings invalidate the positive risk-benefit assessment
- The time schedule and recruitment phase cannot be met, secondary to low recruitment
- In response to recommendations any new findings related to the study device
- The sponsor decides to terminate the study, secondary to device related issues or medical issues

#### 8.3 Termination of a Site by the Study Sponsor

The study may be terminated at an individual investigational site at any time for any of the following reasons below and will be determined by the study sponsor in consultation with site study investigator

- Repeated failure to complete case report forms
- Failure to obtain informed consent
- Failure to report Serious Adverse Events within 24 hours of knowledge
- Loss of or un-accounting of devices
- Repeated protocol violations
- Failure to enroll adequate numbers of patients

#### 8.4 Resumption of Terminated Sites

The sponsor shall not resume the study at a terminated investigational site without prior IRB/EC approval.

#### Protocol amendments

**Amendment 1:** **Protocol version 2 of 24.02.2021.** A more thorough description of the background and rationale behind the study was added under 3.2 and 3.3. Treatment of hemorrhagic stroke**:**

Hematoma evacuation and decompressive craniectomy are options for treating elevated IntraCranial Pressure (ICP) explained by the concept of preventing brain herniation (mechanical damage), reducing ICP, and decreasing the pathophysiological impact of the hematoma on surrounding tissue. Surveillance and monitoring of ICP, Cerebral Perfusion Pressure (CPP), and hemodynamic function. Furthermore, prevention of infection, complications of immobility through positioning and mobilization within physiological tolerance play an important role in optimizing outcomes after ICH. There are multiple approaches to facilitating Cerebrospinal Fluid (CSF) drainage and monitor ICP. Routinely, ICP is measured by use of devices inserted into the brain parenchyma or cerebral ventricles. A Ventricular Catheter (VC) inserted into the lateral ventricle allows for drainage of CSF to help reduce ICP. Although CSF drainage is a vital sequence in patient management, there are reported risks includinginfection and limitations related to erroneous readings associated with current ICP monitors. Physicians lack the appropriate tools to employ active intermittent aspiration and drainage with continuous ICP monitoring.

3.3. Challenges with intraventricular hemorrhage and correlation with clinical outcome

Intraventricular extension of hemorrhage (IVH) is a particularly poor prognostic sign, with expected mortality between 50% and 80%. IVH is a significant and independent contributor to morbidity and mortality, yet therapy directed at ameliorating intraventricular clot has been limited. Conventional therapy centers on managing hypertension and intracranial pressure while correcting coagulopathy and avoiding complications such as rebleeding and hydrocephalus. Surgical therapy alone has not changed the natural history of the disease significantly. Although ventriculostomy appears to be effective in controlling ICP, this technique does little to reduce morbidity and does not address the inflammatory process. The severity of communicating hydrocephalus appears to be related to IVH volume and the duration of exposure of CSF to clotted blood [18–22].

Blood in the ventricular system contributes to morbidity in a variety of ways.

1. Damage to the reticular activating system and thalamus during the acute phase of hemorrhage expansion causes a decreased level of consciousness. Coma appears to be prolonged with both a larger volume of blood in the ventricles and a longer exposure [23, 24].
2. Ventricular blood clots blocking cerebrospinal fluid (CSF) conduits cause acute obstructive hydrocephalus, an immediate life-threatening condition limiting cerebral perfusion and potentially contributing to mass effect and resultant cerebral edema.
3. Blood degradation products become embedded in the arachnoid granulations and may cause permanent occlusion and scarring, inhibiting CSF absorption and causing nonobstructive (communicating) hydrocephalus. Clotted blood may persist in the ventricles for several reasons related to coagulation and fibrinolytic pathways in the ventricular system.
4. The inflammatory reaction caused by blood breakdown products in the ventricles also may affect long-term cognitive function independent of clot volume or mass effect, as demonstrated in experimental IVH [23, 25]. In human IVH, significant cognitive deficits in SAH patients with IVH compared with those without IVH have been found on neuropsychological testing [26].
5. Hematoma growth is an independent determinant of both mortality and functional outcome after ICH [27]. In a secondary analysis of data from the multicenter, randomized, placebo-controlled trial of rFVIIa effectiveness in spontaneous ICH, 170 of 374 patients (45%) had IVH at baseline and 12% (44 of 374) had a greater than 2 mL increase in IVH volume between baseline and 24-hour CT scan [28]. Limiting intraventricular hematoma growth may be an important therapeutic target.

Given the above information, it appears the most important part to treat patients with severe IVH is to remove IVH as soon as possible, dredge cerebrospinal fluid (CSF) circulation, prevent intracranial hypertension, and minimize secondary brain damage. In the traditional treatment, extraventricular drainage can dredge the CSF circulation and reduce the intracranial pressure, but with a high incidence of rebleeding and intracranial infection (Ge et al., 2019; Masoom Abbas, Gopal Varma, Sankar, & Pai, 2019). Therefore, it is always necessary to find a safe and effective treatment for patients with severe IVH in order to quickly eliminate ventricular hemorrhage and prevent recurrent bleeding. With this being stated, there was study performed entitled, Efficacy and safety profile of neuroendoscopic hematoma evacuation combined with intraventricular lavage in severe intraventricular hemorrhage patients *(30)*. The results of this study showed that the hematoma clearance rate in the

**Amendment 2: protocol version 3 of 05.05.21:** Change of randomization fashion from 2:1 to 1:1 due to better funding of the project.

**Amendment 3: Protocol version 4 of 16.01.22:** Change of primary outcome to catheter occlusion from clearance of blood. Further, an inclusion criteria was added: Graeb score of 3 or above. A new sample size calculation and change in no. of intended participants from 104 to 58. Interim analysis should be performed after 20 patients.

**Amendment 4: Protocol version 5 of 28.03.22:** A description of SAE and DMC was added:

For early safety monitoring, an interim analysis is done when 20 subjects have been enrolled in the study (10 subjects per arm). The analyzes will be performed in an intent-to treat fashion.

Monitored safety outcomes are mortality as well as the total number of adverse and serious adverse events related to catheter treatment (infections, bleedings in relation to intervention, displacements of catheters, catheter misplacement). For p-values below 20% early stop of the study will be considered (non-binding) otherwise the study continues until the study end.

Further an Independent Data Monitoring Committee (DMC) has been created with the main purpose of patient safety. The DMC will achieve this by monitoring especially adverse events and severe adverse events and further analyzing the benet vs risk ratio of the treatment. The DMC will evaluate safety of the study after enrollment of 10 patients. The safety evaluation will be based on severe adverse events in each group. For p-values below 20% early stop of the study will be considered (non-binding). Additionally, the DMC will provide an independent scientic review of the interim analysis and recommend continuation or discontinuation of the trial. If the trial passes interim analysis, the DMC will review the nal data as well. The DMC will serve in an advisory capacity to the sponsor.

**Amendment 5: Protocol version 6 of 21.11.22: A section regarding CT fluoroscopy was added:**

It is currently unknown to which extent IRRAflow is able to exchange CSF and perfuse saline in the more distal parts of the ventricular system and subarachnoid space far away for the catheter tip, e.g. the contralateral, 3^rd^ and 4^th^ ventricles, the basal cisterns and sulci, to facilitate wash-out of blood from these spaces and apply medicinal products.

To shed light on this problem, we will evaluate the distribution of the irrigating saline in the ventricles by administrating contrast in the perfusion saline (2 ml Omnipaque, approved for clinical use incl. ventriculography (maximal tolerable dose, 15ml)). During contrast administration, the patient will undergo a fluoroscopy in which real time images will be recorded to visualize the distribution of contrast in the ventricular system of the brain. Various settings of IRRAflow will be explored including different perfusion rates, drainage gradient pressures, etc. We will conduct a series of wash-in and wash-out images to visualize the contrast distribution over a suitable period of time, typically in the range of 30-60 minutes. We will apply a maximum of 10 mSv to each patient for this purpose.

Fluoroscopy is an effective and low-risk imaging tool. Fluoroscopy employs continuous low-dose pulsed radiation with real-time image reconstruction and display, where a normal series (no subtraction) would give a dose of approximately 0,01 mSv. A subtracted series would give 0,4 mSv.
It generally increases patient radiation dose compared with conventional CT scans. A standard head CT applies 2,5 mSv to the patient. However, the peak skin dose from CT fluoroscopy– guided procedures may reach those from other interventional procedures (35). To date, reports of deterministic effects due to CT are extremely rare and have been associated with the combination of repeated multidetector CT studies and fluoroscopic procedures in the same anatomic area (35).

**Experimental outline**

A subgroup of patients will be enrolled from the interventional arm of the ACTIVE study. Only patients stable for transportations and in a good clinical condition will have the fluoroscopy performed.

A baseline 3D fluoroscopy scan will be conducted before Omnipaque is administrated in the irrigating saline. The baseline CT fluoroscopy will apply 0.25 mSv of radiation to the patient. After the baseline CT fluoroscopy, Omnipaque contrast will be administrated in the irrigating saline. Dynamic (live) fluoroscopy will be used to obtain real time video images of the immediate distribution when a bolus of saline is administered. Fluoroscopy (approximately 0,01 mSv per series) may be applied with appropriate intervals to visualize the dynamics of contrast distribution at different time points.

To evaluate the distribution of contrast applied in the irrigation saline over time (wash in), a series of 3D fluoroscopy scans will be performed. A 3D CT fluoroscopy applies 0.2 mSv pr series. The same series of 3D fluoroscopy will be applied to evaluate wash out of the Omnipaque contrast.

In total we expect to apply approximately 3 mSv pr patient. The total amount of radiation will in all cases be significantly below 10 mSv. The majority of the patients in the study are 60-80 years and according to the radiation guideline (from The Central Denmark Region Committee for Health Research Ethics) lifetime risk of cancer therefore very much below the threshold for group 2b.

**Rationale of fluoroscopy**

The fluoroscopy study will provide important information that will be significant to optimize each patient's treatment, specifically to guide the settings of the IRRAflow system to increase perfusion in the relevant areas and understand potential limitations in each case. Furthermore, it will provide crucial information to guide the future use of IRRAflow in the treatment of IVH patients, including optimization of catheter placement for various IVH locations and hemorrhage distributions as well as improve device settings in general. It will provide important insights into the potential therapeutic reach of IRRAflow, and thereby give physicians a better understanding of the areas in the brain, e.g. distal subarachnoid spaces, that can be reached with clot dissolving medication, antibiotic drugs or similar. The information provided by CT fluoroscopy thereby benefits society by means of expected improvement of the IRRAflow treatment and the treatment of IVH in general. We expect this optimized treatment will ultimately prevent secondary brain damage and hydrocephalus and improve neurological outcome and survival by means of accelerated hemorrhage wash-out.
