## Supplementary material for "Safety, harm, and efficacy of IRRAflow^®^ versus external ventricular drainage for intraventricular hemorrhage: A randomized clinical trial": All supplemenatry: Supplementary Material 2.docx

**Supplementary Material 2.**  The IRRAflow® system

IRRAflow® is an advanced drainage system that combines gravity-driven drainage with periodic irrigation and intracranial pressure (ICP) monitoring. The system operates in cycles with the length of each cycle depending on the device settings. Each cycle begins with a one-second irrigation phase, followed by a nine-second ICP measurement phase and ends with a variable-length aspiration phase (10-170 seconds). The control unit offers multiple device settings, allowing users to adjust the irrigation rate (20 to 180 ml/hour) by changing the duration of the aspiration phase or the bolus volume (1 ml or 0.5 ml).

**Irrigation**

IRRAflow® uses a peristaltic bolus infusion mechanism to deliver 0.5 or 1 ml of irrigation over one second. Users can set cumulative irrigation rates between 20 ml/hour and 180 ml/hour. The system is closed but requires a manual change of the 1L Ringer Acetate saline bag when empty. To maintain sterility, it is important to use proper technique when changing the bag, which may need to be done multiple times in every 24-hour period. The system includes an intrinsic ICP monitor for supervisory control of irrigation and aspiration, and automatically deactivates if ICP exceeds a user-defined threshold called “Drain Above”.

**ICP monitoring**

IRRAflow® monitors ICP using a pressure sensor located in the cassette of the device, connected to the irrigation line of the IRRAflow® Tube Set. The sensor is situated proximal to the catheter tip, unlike a standard parenchymal ICP monitor which is located in the brain. The sensor measures ICP as a hydrostatic pressure, so it must be placed in a liquid environment to function properly. The pressure is equivalent to the catheter tip when the irrigation line is fluid-filled, unobstructed, and still. Reliable ICP measurements are critical for irrigation and aspiration operation of the device, which is under intrinsic supervisory control. The IRRAflow® also includes ICP alarm limits to alert users of deviations from the preset limits. The control unit discontinues irrigation and aspiration when an alarm is triggered.

**Aspiration**

Like a standard EVD, IRRAflow® drainage is gravity driven. However, with a standard EVD, the user adjusts the level of the drainage line relative to the catheter tip to define the pressure gradient driving aspiration. Typically, the drain line is placed 5-20 mmHg above the tip of the catheter, so drainage occurs only when the ICP exceeds this level. With IRRAflow®, drainage is regulated by several factors, including the "Drain Above" setting that defines the lower pressure limit for active irrigation, including drainage. When the ICP is within the operational range, drainage is driven by the gradient between the ICP and the level of the drainage bag, which can be set from -15 cmH2O to -103 cmH2O below the catheter tip. This results in active suction of CSF during drainage with gradients ranging from 0 to 75 mmHg.

**Disposables and surgical procedure**

The IRRAflow® system consists of a control unit connected to a tube set, which, in turn, is connected to a bag of saline, a ventricular catheter, and a drainage bag (Figure 1). The catheter is a dual-lumen catheter with an outer diameter of 9fr and two inner lumens, one for irrigation and one for drainage, with an inner diameter of 1.5mm.

To place the catheter, reverse tunneling is performed from a secondary incision behind the burr hole incision (See figure below). The catheter is then fixed in place using a fixation cap and sutures. Unlike some other EVDs, the IRRAflow® catheter does not contain a metal enforcement, which precludes bolted catheter placement. Additionally, the catheter is not coated with antibiotics or silver.

Due to the length of the catheter and the pre-attached male and female luer lock connectors, the standard stylet for neuronavigation is not applicable for use inside the catheter. However, the catheter can be placed using the included stylet provided by the company or in a canal previously made by another neuronavigation-compatible catheter. Alternatively, the catheter can be placed with the stylet from the neuronavigation placed in the side-hole of the tip of the catheter.

**Differences between IRRAflow® and Silverline**® **EVD**

1.     Irrigation versus no irrigation.

2.     Tunneled catheter versus bolted catheter.

3.     Uncoated catheter versus coated catheter.

4.     Drainage dependent on ICP measurement in device versus standard passive drainage.

5.     Change of saline bag for irrigation versus no change of saline bag.

6.     Active aspiration (negative pressure gradient versus passive drainage

7.     Inner catheter diameter: 1.5 mm (outer diameter: 9 French) versus 1.9 mm (outer diameter: 10 French)

8.     Navigation with stylet in side-hole versus navigation stylet inside EVD as intended

Due to the intent-to-treat analysis method used in the study, the subgroup of participants in the intervention group who were treated with bilateral drains were assigned to the intervention group regardless of whether the AE could be ascribed to IRRAflow® or the contralateral EVD.

**Differences between IRRAflow**® **and EVD placement and fixation**

The IRRAflow® catheter was introduced using a reverse tunneling technique and secured with a sutured fixation cap, whereas the Silverline® catheter was placed with a minimally invasive skin incision and bolt fixation. Differences in the surgical procedure between the intervention and the control are shown in the figure below. A shows the reverse-tunneled IRRAflow® catheter, B shows placement of the IRRAflow® catheter in the ventricular system, and C shows the completed surgery with the attached IRRAflow® catheter and the skin sutured (intervention). D. shows the bolt-connected Silverline® catheter (control)

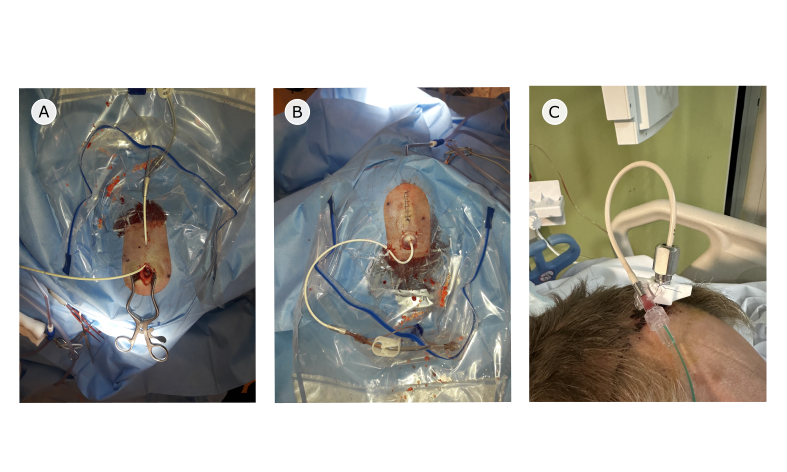
