## Supplementary material for "Safety, harm, and efficacy of IRRAflow^®^ versus external ventricular drainage for intraventricular hemorrhage: A randomized clinical trial": All supplemenatry: Supplementary Material 3.docx

**Supplementary Material 3.** Standard operation procedure (SOP), IRRAflow® treatment in the Active Study.

**Introduction and purpose**

This SOP describes the IRRAflow® treatment in the Active Study. The purpose is to align placement and treatment with the IRRAflow® in the ACTIVE Study

**Catheter placement**

1.   The IRRAflow® catheter should be placed by reverse tunneling and not on a bolt. The catheter should be tunneled from a minimal lateral incision towards a burr hole incision at the drain entry point. The catheter is reverse tunneled after the burr hole is made, the dura opened, and a corticotomy is performed. The catheter is then inserted into the lateral ventricle. The catheter can either be placed 1) without neuronavigation using the included stylet belonging to the catheter, 2) or in a canal made prior to insertion using another neuronavigation-compatible catheter, or 3) with a neuronavigation stylet in the side-hole of the catheter tip. Upon catheter placement, correct position is verified by steady flow of CSF from the catheter. After surgery, a head computed tomography (CT) should be performed before irrigation is initiated to ensure correct catheter positioning in the ventricular system.

2.   All participants should have a Raumedic Neurovent-P ICP monitor placed in the same hemisphere as the IRRAflow® catheter. ICP should be evaluated from the Raumedic device using an MPR 1 or 2 datalogger.

**Standard IRRAflow® settings**

1.   Alarm limits: Low -5 mmHg, High 35 mmHg. ICP is monitored on the Raumedic device, and the participant is treated according to standard guidelines regarding ICP limits. In most cases, ICP must be kept below 20 mmHg.

2.   Bolus: 1 ml

3.   Drain above: 10 mmHg under consideration of the participants’ clinical condition, ICP, potential untreated aneurysm, etc.

4.   Irrigation rates in prioritized order:

a.     High (180 ml/hour): Cycle time 20 sec., bag level 29 cm

b.     Medium (90 ml/hour): Cycle time 40 sec., bag level 21 cm

c.      Low (20 ml/hour): Cycle time 180 sec., bag level 15 cm

Medium and low device settings are used in participants with marginal ICP control.

5.     Ringer Acetate, 1 L bags, room temperature is used for irrigation.

**Participant observation**

1.   The participant should always be observed in the neurointensive care unit.

2.   IRRAflow® is set at “drain only” mode if ICP values exceed > 25 mmHg measured at the Raumedic device.

3.   All participants shall have head CTs at days 0, 2, 4, 6 and 8.

**End of IRRAflow**® **treatment:**

1.   IRRAflow® irrigation line should be closed and disconnected.

2.   The drainage line should be disconnected under sterile conditions and attached to a standard drainage bag.

3.   The participant may be moved to the neurosurgical ward and the drain treatment may be set to continue as for standard EVD treatment.
