## Supplementary material for "Safety, harm, and efficacy of IRRAflow^®^ versus external ventricular drainage for intraventricular hemorrhage: A randomized clinical trial": All supplemenatry: Supplementary Material 4.docx

**Supplementary Material 4** SAEs with possible relation to intervention and/or catheter treatment.

The following specific conditions were considered *a priori* to be SAEs with a potential causal relationship to intervention or catheter treatment:

- Death
- CNS infections: e.g., ventriculitis in relation to catheter treatment
- Intracranial hemorrhage occurring after inclusion: e.g., hemorrhage in relation to placement of the catheter, treatment with catheter, or removal of catheter.
- Catheter displacements: catheter tip displacement evaluated on control CT. Displacements with catheter tip in parenchyma where irrigation or medicine administration has appeared was categorized as a SAE)
- Catheter misplacement: A control head CT was performed after catheter placement. Catheters located in an incorrect place were categorized as misplaced.
- Catheter occlusion leading to surgical replacement of the catheter or administration of alteplase to decrease the risks of severe hydrocephalus.

CNS = Cerebrospinal liquids
CT =Computed tomography
SAE = Severe adverse event
