## Supplementary material for "Safety, harm, and efficacy of IRRAflow^®^ versus external ventricular drainage for intraventricular hemorrhage: A randomized clinical trial": All supplemenatry: Supplementary Material 5.docx

**Supplementary Material 5.** Time-to-catheter occlusions shown as cumulative incidence plots for catheter occlusions. A shows the cumulative incidence for all catheters (hazard ratio 4.4 (95%CI 0.6-31.2), p=0.141 and B for the primary catheter only (hazard ratio 3.2 (95%CI 0.35-28.7), respectively. Red line: cumulative incidence IRRAflow®, blue line: cumulative incidence EVD. Dotted lines: 95%CI for each cumulative incidence plot.

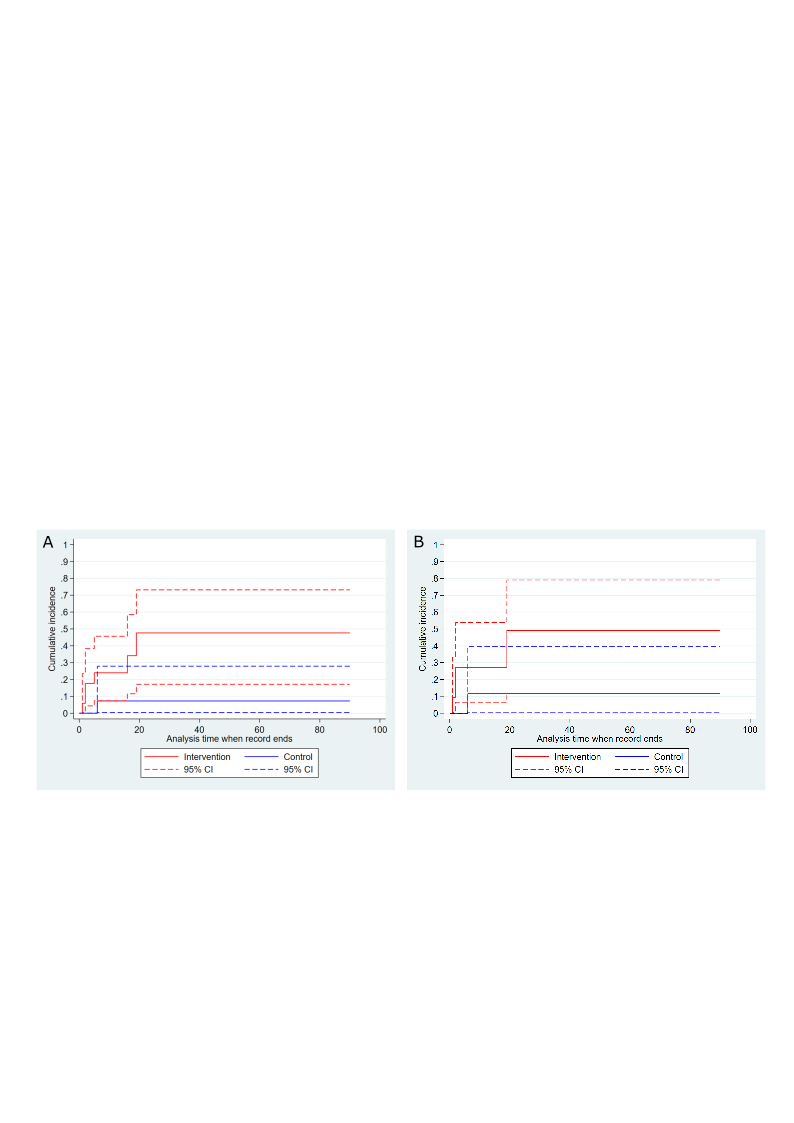
