## Supplementary material for "Safety, harm, and efficacy of IRRAflow^®^ versus external ventricular drainage for intraventricular hemorrhage: A randomized clinical trial": All supplemenatry: Supplementary Material 6.docx

**Supplementary Material 6.** The full DMC safety report and interim analysis.

Interim analysis results presented for the DMC for the first 20 patients:

**Risk differences and incidence rates of SAEs**

|  | **Intervention (n = 10)** | | **Control (n = 10)** | |  |  |
| --- | --- | --- | --- | --- | --- | --- |
| **Risk differences** | | | | | | |
|  | **n** | **%** | **n** | **%** | **Risk difference** | **p-value** |
| Risk difference of experiencing ≥ 1 SAE | 7 | 70% | 2 | 20% | 0.5 95%CI (0.123-0.877) | **0.02** |
| **Incidence rates** | | | | | |  |
|  | **Incidence rate** |  | **Incidence rate** |  | **Incidence rate ratio** | **p-value** |
| Incidence rate of experiencing  ≥ 1 SAE, excluding death* | 0.009 |  | 0.004 |  | 2.68 95%CI (0.6-12.9) | **0.2** |
| Incidence rate in number of SAEs divided by person time, excluding death* | 0.026 |  | 0.012 |  | 6.0 95%CI (1.4-26.6) | **0.012** |

Statistically significant findings (alpha 0.2) are marked in bold.

*Death was excluded as death was not causally related to either the intervention or to catheter treatment

**Severe adverse event timeline**.

All SAEs plotted in a timeline from inclusion to follow-up for the first 20 patients at the time of the interim analysis. Super-stitched numbers represent the grade of the observed SAE. Furthermore, occlusion of primary and secondary catheters is plotted. The number of AEs is significantly higher in the intervention group (incidence rate ratio = 6.0 95%CI (1.4-26.6), p=0.012)

CI = Confidence interval
DMC = Data safety monitoring committee
SAE = Severe adverse events

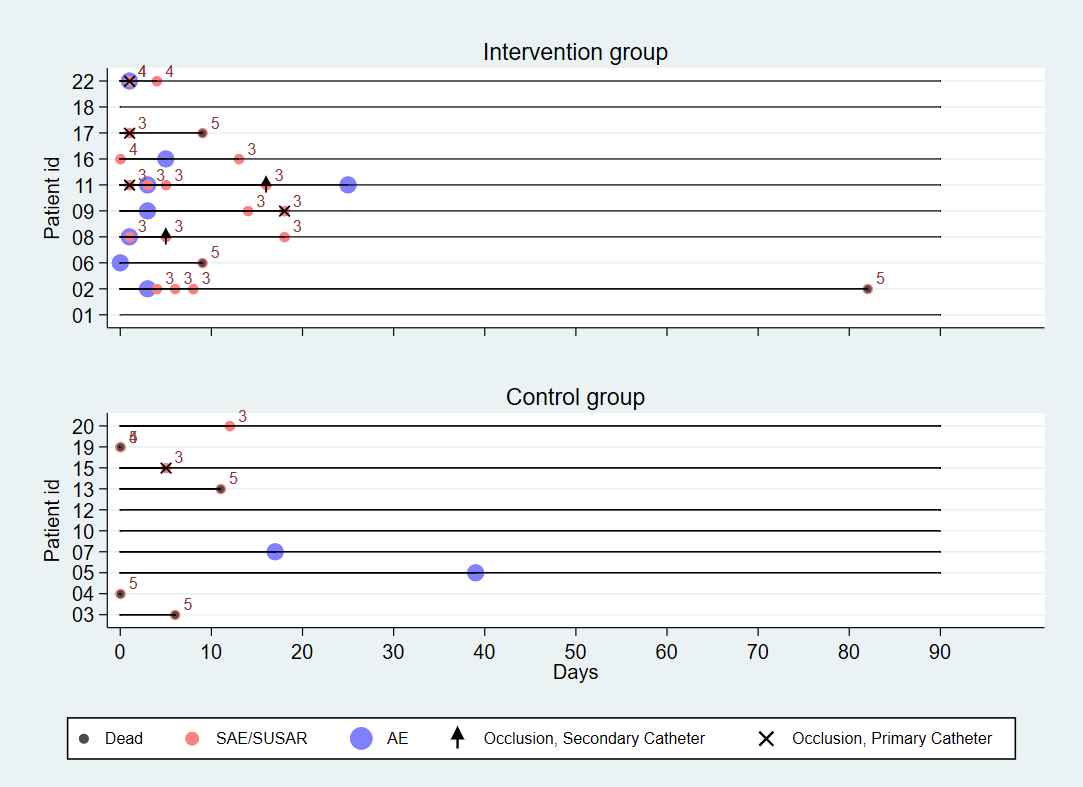

AE = Adverse events
SAE = Severe adverse events
SUSAR = Suspected unexpected serious adverse reaction

**Causality assessment of all SAEs**

Detailed table of all severe adverse events (SAEs) in the intervention and control group. The WHO-UMC classification of the different SAEs is shown along with the assumed cause of the SAE based on the root cause analysis.
