## Supplementary material for "Safety, harm, and efficacy of IRRAflow^®^ versus external ventricular drainage for intraventricular hemorrhage: A randomized clinical trial": All supplemenatry: Supplementary Material 8.docx

**Supplementary Material 8.** Frequencies of adverse events. The table shows the numbers, frequencies, and grade of all AEs. A patient is registered as having an AE if the AE occurred at least once, regardless of severity. Patients are divided into those treated with IRRAflow® (left column) and those treated with EVD (middle column). The right column shows results for all patients collectively.

| Type of AE |  | | | | | | | | |
| --- | --- | --- | --- | --- | --- | --- | --- | --- | --- |
|  | Intervention (n = 11) | | | Control (n = 10) | | | Total (n = 21) | | |
|  | n | % | 95%CI | n | % | 95%CI | n | % | 95%CI |
| Catheter displacements |  |  |  |  |  |  |  |  |  |
| Grade 1  Grade 2  Grade 3  Grade 4  Grade 5 | -  2  3  -  - | 18.2%  27.3% | (2.3- 51.8)  (6.0-60.9) | -  1  -  -  - | 10% | (0.2-44.5) | -  3  3  -  - | 14.3%  14.3% | (3.0-36.3)  (3.0-36.3) |
| Infections |  |  |  |  |  |  |  |  |  |
| Grade 1  Grade 2  Grade 3  Grade 4  Grade 5 | -  -  3  -  - | 27.3% | (6.0-60.9) | -  -  1  -  - | 10% | (0.2-44.5) | -  -  4  -  - | 19.1% | (5.4-41.9) |
| Catheter replacements |  |  |  |  |  |  |  |  |  |
| Grade 1  Grade 2  Grade 3  Grade 4  Grade 5 | -  -  4  -  - | 36.4 % | (10.9-69.2) | -  -  1  -  - | 10% | (0.2-44.5) | -  -  5  -  - | 23.8% | (8.2-47.2) |
| Rebleeding |  |  |  |  |  |  |  |  |  |
| Grade 1  Grade 2  Grade 3  Grade 4  Grade 5 | 1  -  -  2  - | 9.1%  18.2% | (0.2-41.3)  (2.2-51.8) | 1  -  -  -  - | 10% | (0.2-44.5) | 2  -  -  2  - | 9.5%  9.5% | (1.1-30.3)  (1.1-30.3) |
| Disconnect of tubesystem |  |  |  |  |  |  |  |  |  |
| Grade 1  Grade 2  Grade 3  Grade 4  Grade 5 | 1  -  -  -  - | 9.1% | (0.2-41.3) | 1  -  -  -  - | 10% | (0.2-44.5) | 2  -  -  -  - | 9.5% | (1.1-30.3) |
| CSF leakage |  |  |  |  |  |  |  |  |  |
| Grade 1  Grade 2  Grade 3  Grade 4  Grade 5 | -  4  -  -  - | 36.4% | (10.9-69.2) | -  -  -  -  - |  |  | -  4  -  -  - | 19.1% | (5.4-41.9) |
| Died due to initial poor prognosis |  |  |  |  |  |  |  |  |  |
| Grade 5 | 4 | 36.4% | (10.9-69.2) | 4 | 40% | (12.6-73.8) | 8 | 38.1% | (18.1-61.5) |
