## Supplementary material for "Safety, harm, and efficacy of IRRAflow^®^ versus external ventricular drainage for intraventricular hemorrhage: A randomized clinical trial": All supplemenatry: Supplementary Material 9.docx

**Supplementary Material 9.** Causality of adverse events

AEs related to catheter treatment

|  | Related to catheter treatment | | | | |
| --- | --- | --- | --- | --- | --- |
|  | Intervention | Control | Total | | |
| **Type of AE** | n | n | n | % | 95%CI |
| Catheter displacement | 6 | 1 | 7 | 23.33 | (9.9-43.3) |
| Infections | 3 | 1 | 4 | 13.33 | (3.8-30.7) |
| Catheter replacements | 7 | 1 | 8 | 26.67 | (12.3-45.9) |
| Rebleeding | 2 | 0 | 2 | 6.67 | (0.8-22.1) |
| Disconnect of tube system | 1 | 1 | 2 | 6.67 | (0.8-22.1) |
| CSF leakage | 4 | 0 | 4 | 13.33 | (3.8-30.7) |

AEs related to different aspects of the IRRAflow® system

|  | Related to intervention | | | | | | | | | | | | | | |
| --- | --- | --- | --- | --- | --- | --- | --- | --- | --- | --- | --- | --- | --- | --- | --- |
|  | Irrigation (vs no irrigation) | | | | | Tunneled (vs bolted) placement | | | | | Uncoated catheter (vs coated catheter) | | | | |
|  | Intervention | Control | Total | | | Intervention | Control | Total | | | Intervention | Control | Total | | |
| **Type of AE** | n | n | n | % | 95%CI | n | n | n | % | 95%CI | n | n | n | % | 95%CI |
| Catheter displacement | 0 | 0 | 0 | 0 |  | 6 | 0 | 6 | 20,00 | (7.7-38.6) | 0 | 0 | 0 | 0 |  |
| Infections | 0 | 0 | 0 | 0 |  | 3 | 0 | 3 | 10,00 | (2.1-26.5) | 3 | 0 | 3 | 10,00 | (2.1-26.5) |
| Catheter replacements | 0 | 0 | 0 | 0 |  | 1 | 0 | 1 | 3,33 | 0.08-17.2) | 0 | 0 | 0 | 0 |  |
| Rebleeding | 2 | 0 | 2 | 6,67 | (0.8-22.1) | 0 | 0 | 0 | 0 |  | 0 | 0 | 0 | 0 |  |
| Disconnect of tube system | 0 | 0 | 0 | 0 |  | 0 | 0 | 0 | 0 |  | 0 | 0 | 0 | 0 |  |
| CSF leakage | 0 | 0 | 0 | 0 |  | 4 | 0 | 4 | 13,33 | (3.8-30.7) | 0 | 0 | 0 | 0 |  |
|  | Device ICP dependent vs device ICP independent | | | | | Active suction vs passive drainage | | | | | Catheter inner diameter 1.5 (vs. 1.9) | | | | |
|  | Intervention | Control | Total | | | Intervention | Control | Total | | | Intervention | Control | Total | | |
| **Type of AE** | n | n | n | % | 95%CI | n | n | n | % | 95%CI | n | n | n | % | 95%CI |
| Catheter displacement | 0 | 0 | 0 | 0 |  | 0 | 0 | 0 | 0 |  | 0 | 0 | 0 | 0 |  |
| Infections | 0 | 0 | 0 | 0 |  | 0 | 0 | 0 | 0 |  | 0 | 0 | 0 | 0 |  |
| Catheter replacements | 4 | 0 | 4 | 13,33 | (3.8-30.7) | 4 | 0 | 4 | 13,33 | (3.8-30.7) | 4 | 0 | 4 | 13,33 | (3.8-30.7) |
| Rebleeding | 0 | 0 | 0 | 0 |  | 0 | 0 | 0 | 0 |  | 0 | 0 | 0 | 0 |  |
| Disconnect of tube system | 0 | 0 | 0 | 0 |  | 0 | 0 | 0 | 0 |  | 0 | 0 | 0 | 0 |  |
| CSF leakage | 0 | 0 | 0 | 0 |  | 0 | 0 | 0 | 0 |  | 0 | 0 | 0 | 0 |  |
