## Supplementary material for "Safety, harm, and efficacy of IRRAflow^®^ versus external ventricular drainage for intraventricular hemorrhage: A randomized clinical trial": All supplemenatry: Table supplementary 6 SAE overview and causality.docx

|  | Ventriculitis | | Catheter displacements | | Catheter replacements or contra lat. or tPA | | Rebleeding | | Died | |
| --- | --- | --- | --- | --- | --- | --- | --- | --- | --- | --- |
| Id | **Intervention** | **Catheter treatment** | **Intervention** | **Catheter treatment** | **Intervention** | **Catheter treatment** | **Intervention** | **Catheter treatment** | **Intervention** | **Catheter treatment** |
| Id 1 |  |  |  |  |  |  |  |  |  |  |
| Id 2 | I, possible, tunneling, no coating | I, certain | I, probable, irrigation into parenchyma  fixation method  I, probable,  fixation method | I, certain  I, certain |  |  |  |  | I, unlikely | I unlikely |
| Id 6 |  |  |  |  |  |  |  |  | I, unlikely |  |
| Id 8 | I, possible,  tunneling, no coating | I, certain |  |  | I, probable,  displacement and new catheter placed due to fixation method and short tube set of the IRRAflow.  I, unlikely  (Contra lat. EVD occlusion), replacement + tPA due to blood clot | I, certain  I, certain |  |  |  |  |
| Id 9 | I, possible  tunneling, no coating | I, certain |  |  | I, probable  IRRAflow occlusion, inner diameter | I, certain |  |  |  |  |
| Id 11 |  |  | I, probable, irrigation and tPA into parenchyma,  fixation method | I, certain | I, probabale, , IRRAflow occlusion, inner diameter, contra lat. EVD placement  I, probable  IRRAflow occlusion, tPA, inner diameter  I, unlikely  EVD occlusion | I, certain  I, certain  I, certain |  |  |  |  |
| Id 16 |  |  | I, probable, irrigation in to parenchyma  fixation method | I, certain |  |  | I, unlikely (spontaneous new hemorrhage) | I, unlikely | I, Unlikely | I, Unlikely |
| Id 18 |  |  |  |  |  |  |  |  |  |  |
| Id 22 |  |  |  |  | I, probable  IRRAflow occlusion, inner diameter contra lat. EVD placed | I, certain | I, probable,  irrigation  I, probable, irrigation | I, certain  I, certain |  |  |
| Id 23 |  |  |  |  |  |  |  |  | I, unlikely | I, unlikely |

|  | Ventriculitis | | Catheter displacements | | Catheter replacements or contra lat. or tPA | | Rebleeding | |  | |
| --- | --- | --- | --- | --- | --- | --- | --- | --- | --- | --- |
| Id | **Intervention** | **Catheter treatment** | **Intervention** | **Catheter treatment** | **Intervention** | **Catheter treatment** | **Intervention** | **Catheter treatment** | **Intervention** | **Catheter treatment** |
| Id 3 |  |  |  |  |  |  |  |  | I, unlikely | I, unlikely |
| Id 4 |  |  |  |  |  |  |  |  | I, unlikely | I, unlikely |
| Id 5 |  |  |  |  |  |  |  |  |  |  |
| Id 7 |  |  |  |  |  |  |  |  |  |  |
| Id 10 |  |  |  |  |  |  |  |  |  |  |
| Id 12 |  |  |  |  |  |  |  |  |  |  |
| Id 13 |  |  |  |  |  |  |  |  | I, unlikely | I, unlikely |
| Id 15 |  |  |  |  | !, EVD occlusion unlikely | I, certain |  |  |  |  |
| Id 19 |  |  |  |  |  |  | I, unlikely | I, unlikely (coil) | I, unlikely | I, unlikely |
| Id 20 | I, unlikely | I, certain |  |  |  |  |  |  |  |  |
